# Cross-Sectional and Longitudinal Patterns of Atrophy in Thalamic and Deep Gray Matter Nuclei in Frontotemporal Dementia

**DOI:** 10.1101/2025.02.10.25322025

**Authors:** A. Banerjee, F. Yang, J. Dutta, A. Cacciola, M. Hornberger, M. Saranathan

## Abstract

**INTRODUCTION:** Frontotemporal dementia involves progressive atrophy in deep gray matter nuclei, including the thalamus and basal ganglia (such as the caudate, putamen, nucleus accumbens, and globus pallidus), which are critical for cognition and behavior. This study examined cross-sectional and longitudinal atrophy using a state-of-the-art multi-atlas segmentation method sTHOMAS.

**METHODS:** T1-weighted MRI scans from 274 participants at baseline and 237 at follow-up obtained from the Frontotemporal Lobar Degeneration Neuroimaging Initiative database were analyzed using sTHOMAS. Group differences were assessed using ANCOVA, adjusting for age, gender and intracranial volume as covariates.

**RESULTS:** Atrophy was significant in the mediodorsal, pulvinar, anterior ventral nuclei, nucleus accumbens, and claustrum, with bvFTD most affected cross-sectionally. Longitudinally, the nucleus accumbens, mediodorsal, and pulvinar nuclei declined further. Atrophy correlated with naming (mediodorsal), working memory (ventrolateral posterior), and executive dysfunction (nucleus accumbens) neuropsychological tests.

**DISCUSSION:** These findings highlight progressive, nucleus-specific atrophy in FTD and emphasize the importance of cross-sectional as well as longitudinal imaging and sex-specific analyses in understanding disease progression.

## 1. BACKGROUND

While Alzheimer’s disease is the most prevalent dementia, Frontotemporal Dementia (FTD) is a leading cause of early-onset dementia, typically manifesting between ages 45 and 65 [1]. Sporadic FTD encompasses heterogeneous syndromes and are classified into subtypes notably-behavioral variant FTD (bvFTD) and primary progressive aphasias—semantic variant (svFTD) and progressive non-fluent aphasia (PNFA).

BvFTD, the most common subtype, involves profound personality changes, disinhibition, apathy, and ritualistic behaviors, reflecting early frontal lobe degeneration [2–6]. SvFTD is associated with anterior temporal atrophy, impairing word comprehension and naming, while PNFA, linked to left-hemisphere atrophy, leads to halting speech and grammatical errors [7].

The thalamus, a central hub with extensive cortical-subcortical connections, regulates sensory integration, cognition, emotion, and motor coordination [9]. Its nuclei, crucial for sensory processing, attention, and memory, exhibit selective vulnerability in FTD, providing key pathophysiological insights [10,11]. Mediodorsal (MD) nuclei, integral to memory and executive function, consistently atrophy across all FTD subtypes, while pulvinar nuclei, implicated in visual and language functions, show phenotype-specific degeneration, particularly in C9orf72 carriers [12]. The anterior ventral nuclei, critical for behavioral and memory processes, also exhibit significant volume loss [7]. Examining these nuclei individually aids biomarker identification and targeted interventions [13]. The striatum (caudate, putamen, nucleus accumbens) also exhibits distinct atrophy patterns in FTD. The dorsal striatum (caudate, putamen) supports sensorimotor functions, while the ventral striatum (nucleus accumbens) regulates behavior and emotion [14–19]. BvFTD shows pan-striatal atrophy with right-hemisphere dominance linked to behavioral symptoms [20], while PNFA and svFTD display left-predominant atrophy in language-related regions.

Conventional T1-weighted MRI lacks the contrast needed for precise subcortical segmentation, particularly for thalamic nuclei [21,22]. Advanced methods like White-Matter-nulled (WMn) MPRAGE enhance intrathalamic contrast, and THalamus Optimized Multi Atlas Segmentation (THOMAS) [23,24] leverages this improved contrast. However, specialized WMn sequences are absent from clinical databases such as Alzheimer’s Disease Neuroimaging Initiative (ADNI) and the Laboratory of Neuro Imaging (LONI).

To address these limitations, the Histogram-based Polynomial Synthesis (HIPS) technique was developed, leading to HIPS-THOMAS, which first synthesizes WMn-like contrast from standard T1-weighted images prior to improved segmentation of thalamic nuclei and basal ganglia [25,26]. Recently, HIPS-THOMAS was extended to segment additional deep gray matter structures, including the caudate, putamen, nucleus accumbens, globus pallidus (internal and external), claustrum, and red nucleus, enabling comprehensive subcortical analyses. (more information in **Table 1**).

**Table 1.**
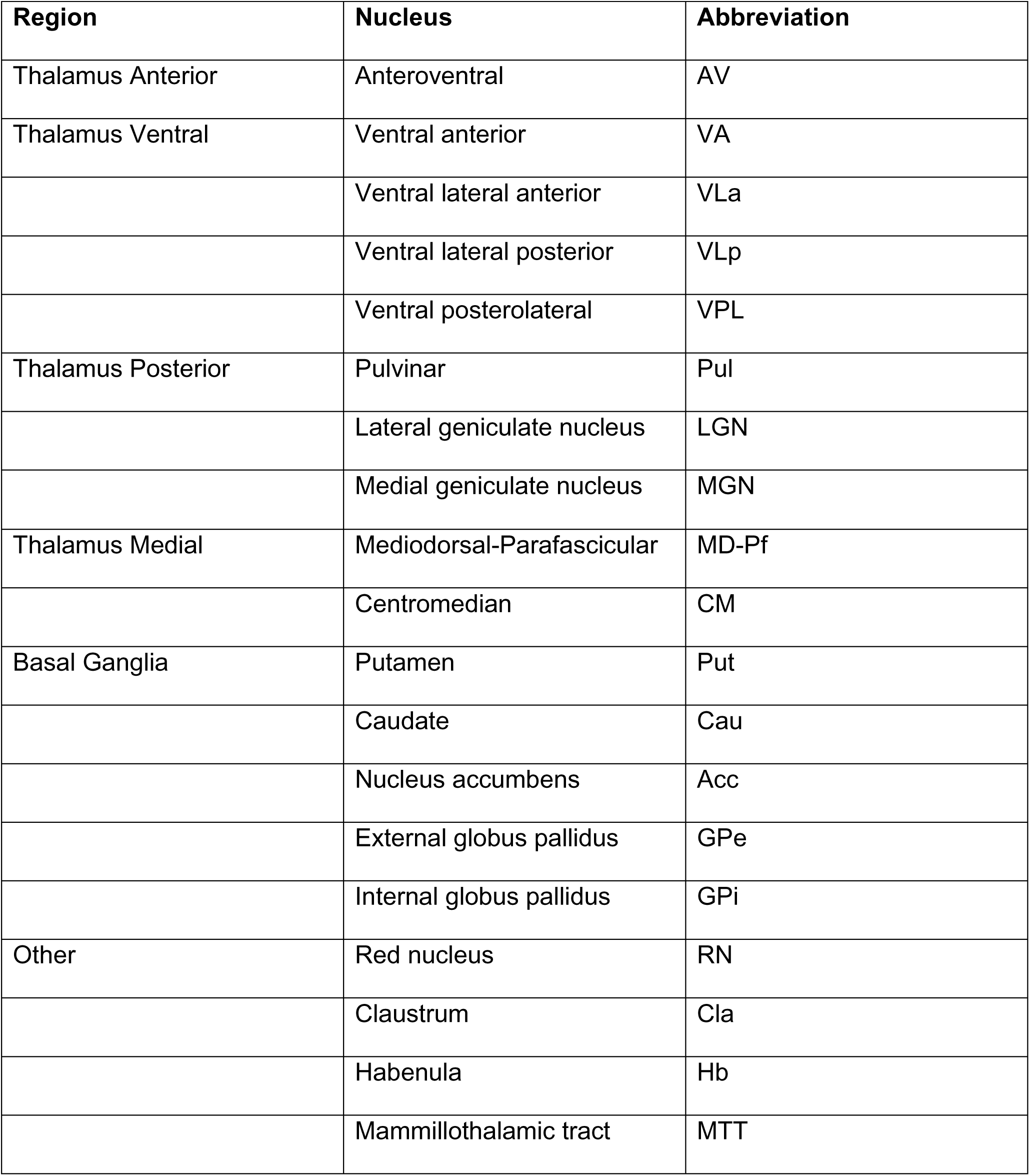
Regions and nuclei segmented by sTHOMAS using the nomenclature established by Morel. [43]

In this study, we applied this novel deep gray nuclei segmentation method (called sTHOMAS) to T1-weighted MRI scans from the LONI FTLDNI database to characterize cross-sectional and longitudinal atrophy patterns, examine gender differences, and evaluate correlations with neuropsychological test scores. Given the variability in FTD progression, biological sex is a key factor shaping neurodegeneration, with studies in bvFTD showing that women exhibit greater frontotemporal atrophy despite similar clinical symptoms, along with greater cognitive decline, while men display more pronounced behavioral disturbances. [27,28]. Given these findings, examining sex differences in deep gray matter atrophy is essential to identifying disease-specific vulnerabilities. Building on from these previous findings and addressing gaps in the literature, we hypothesize that sTHOMAS will enhance the precise delineation of deep gray matter structures, allowing for the identification of subtype-specific atrophy patterns in FTD. Specifically, we anticipate progressive degeneration in key nuclei, including the MD, AV, pulvinar, nucleus accumbens, and putamen.

## 2. METHODS

### 2.1. Data Acquisition

Data for this study were obtained from the Frontotemporal Lobar Degeneration Neuroimaging Initiative (FTLDNI) database (NIH Grant R01 AG032306). Funded by the National Institute on Aging and coordinated through the UCSF Memory and Aging Center, FTLDNI aimed to develop neuroimaging and analytical tools for tracking frontotemporal lobar degeneration (FTLD). Data collection involved collaborative efforts across multiple sites in the United States, including University of California at San Francisco (UCSF), Massachusetts General Hospital (MGH), and Mayo Clinic. High-resolution T1-weighted MRI scans were acquired using 3 Tesla scanners. Institutional Review Boards at all participating institutions approved the study protocols, and informed consent was obtained. This study utilized timepoint 1 data downloaded from the LONI platform in November 2023 and timepoint 2 data downloaded in April 2024.

### 2.2. Imaging and Segmentation

T1-weighted MRI scans were obtained using 3D MPRAGE sequences or equivalent protocols with consistent high-resolution settings across sites (1–1.2 mm isotropic voxel size). Thalamic nuclei segmentation was performed using the sTHOMAS method, an adaptation of the THOMAS pipeline [24] [25]. This pipeline enhances intrathalamic contrast through a Histogram-Based Polynomial Synthesis (HIPS) preprocessing step, simulating WMn imaging contrast. Segmentation utilized multi-atlas labeling and intensity normalization, enabling accurate delineation of thalamic subregions from conventional T1-weighted images. Intracranial volume (ICV) values obtained using FreeSurfer (recon-all command, eTIV) were used to adjust for variability in head size and morphology in analyses. The segmentation process and regional labeling of thalamic and other deep gray matter nuclei using sTHOMAS, showcasing representative T1-weighted and white matter nulled images, along with segmented overlays highlighting key subcortical regions is shown in **Figure 1**.

**Figure 1.**
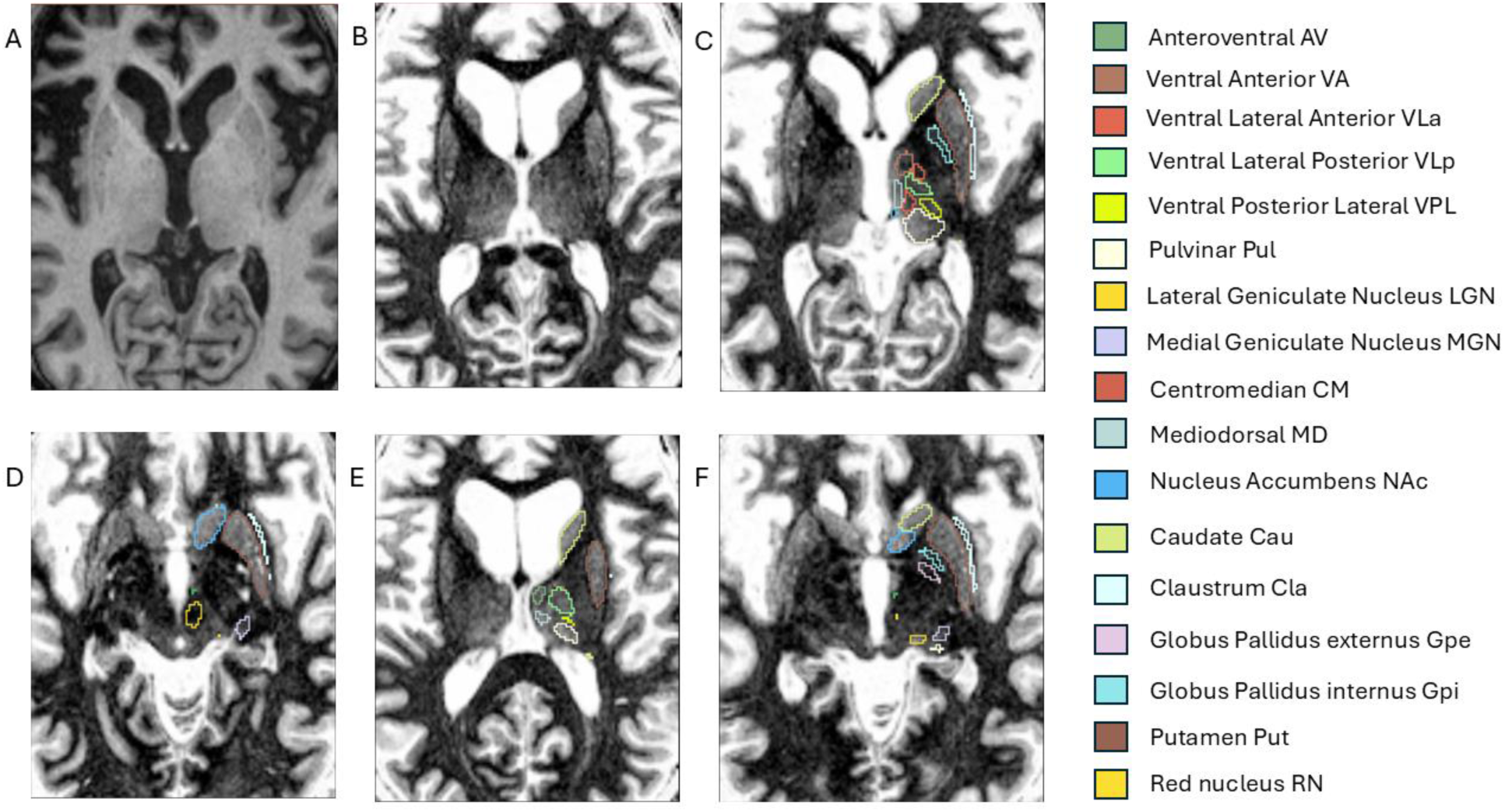
A representative Behavioral variant Frontotemporal Dementia (bvFTD) subject showing cropped T1 (**A**) and HIPS-synthesized white matter nulled (WMn) image (**B**) **(C-F)** Thalamic and deep gray matter nuclei segmentation outputs from sTHOMAS overlaid in the left hemisphere, showing regions including Ventral Anterior (VA), Ventral Lateral Anterior (VLa), Ventral Lateral Posterior (VLp), Ventral Posterior Lateral (VPL), Pulvinar (Pul), Centromedian (CM), Mediodorsal (MD), Caudate (Cau), Claustrum (Cla), Globus Pallidus Internus (Gpi), and Putamen (Put), Nucleus Accumbens (NAc), Red Nucleus (RN), the Anteroventral Nucleus (AV), Globus Pallidus Externus (Gpe), Lateral Geniculate Nucleus (LGN), and Medial Geniculate Nucleus (MGN).

### 2.3. Data Analyses

#### 2.3.1. Cross-sectional data

Group differences in thalamic and other deep gray matter nuclei volumes were assessed using analysis of covariance (ANCOVA), adjusting for age, sex, and ICV, followed by Dunnett’s test for post hoc comparisons against the control group. Effect sizes were calculated using Cohen’s d. Normality of data distributions was verified using the Shapiro-Wilk test. Gender-specific analyses and longitudinal median atrophy rates between timepoint 1 (baseline) and timepoint 2 (follow-up) were calculated to capture patterns of progression and variability across groups.

Outlier detection was performed for all thalamic nuclei, basal ganglia (caudate nucleus, putamen, internal and external globus pallidus, and nucleus accumbens), claustrum, and red nucleus volumes, using the 2×IQR rule. For each nucleus and diagnostic group, the interquartile range (IQR) was computed, and volumes falling outside two IQRs from the first or third quartile were flagged as outliers. Detected outliers were blanked out to prevent undue influence on statistical analyses. Being slightly more conservative than the usual 1.5×IQR, we opted for the 2×IQR to avoid undue reduction of sample size.

Adjusted mean volumes **(Supplementary Table 1)** were computed for each group using estimated marginal means derived from the ANCOVA model. To quantify differences between patient groups and controls, we calculated the percentage reduction in adjusted volumes relative to controls (**Table 3 shows values from timepoint 1 and Supplementary Table 2 shows values from timepoint 2**). To control for potential confounding factors, all volumes were adjusted for ICV, age, and sex using the following adjustment formula:

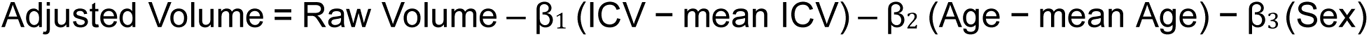

#### 2.3.2. Longitudinal data

We calculated the percentage of median atrophy in thalamic and other deep gray matter nuclei relative to baseline volumes for each subject. The time intervals between scans varied, ranging from 18 to 36 months, depending on data availability. When a 24 month follow-up was available, it was prioritized; otherwise, the closest available time points were used. Atrophy rates were derived as: 100*((Baseline Volume−Follow-up Volume)/ Baseline Volume × Time Interval (years)). To assess group-wise differences, a one-way ANOVA was performed to evaluate whether atrophy varied significantly across clinical groups (CN, bvFTD, PNFA, and svFTD), with CN as the reference. Post hoc Dunnett’s tests were conducted to identify pairwise differences between each clinical group and CN, with adjusted p-values (*p < 0.05, **p < 0.01, ***p < 0.001) to account for multiple comparisons.

### 2.4. Partial Correlation Analysis

#### Adjustment of Deep Gray Nuclei Volumes for Neuropsychological Analysis

Deep gray matter volumes including thalamic nuclei volumes were adjusted for individual ICV to ensure comparability across subjects. Adjusted volumes were calculated using linear regression models with residual volumes normalized to the mean ICV of the cohort. This adjustment accounted for individual variations in brain size, isolating volume changes specific to the nuclei.

The adjusted volume calculation is based on a linear regression model that accounts for the effects of ICV and age. The adjustment is based on how much an individual’s ICV and age deviate from the group means, weighted by the respective beta coefficients derived from the control group.

The following formula was used for calculation:

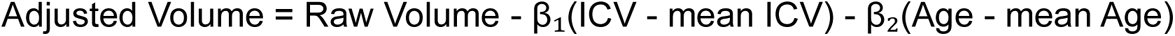

Where:

- Raw Volume is the original volume measurement for a specific thalamic nucleus
- β₁ is the regression coefficient for ICV (obtained from the control group)
- β₂ is the regression coefficient for Age (obtained from the control group)
- mean ICV is the average ICV of the group being adjusted
- mean Age is the average age of the group being adjusted

For each group (bvFTD, PNFA, svFTD), partial correlation analyses were performed to examine the relationships between thalamic and other deep gray nuclei volumes and neuropsychological test scores. To isolate nuclei-specific contributions, correlations were computed while controlling for the contribution of the whole thalamus (both left and right), ensuring that the observed relationships reflected specific nuclei-level changes rather than overall thalamic volume effects. Bonferroni correction was applied to account for multiple comparisons, with a significance threshold of 𝑝 < 0.05 / 8, corresponding to the inclusion of eight prioritized structures.

The prioritized structures included the AV nucleus, MD nucleus, VLp nucleus, pulvinar nucleus, nucleus accumbens, caudate, putamen, and claustrum. These structures were selected based on their high effect sizes (Cohen’s d) and significant percentage changes in median atrophy between bvFTD, PNFA, svFTD, and CN groups. These metrics highlighted their potential as key regions of interest in understanding disease-specific structural alterations. Correlations were annotated with significance levels (* for 𝑝 < 0.05, ** for 𝑝 < 0.01, and *** for 𝑝 < 0.001) to emphasize the most critical findings.

#### 2.4.1. Neuropsychological Tests

1. **Boston Naming Test (15-item)**:

◦ Measure: Total score
◦ Abbreviation: BNTCORR
2. **Modified Trails:**

◦ Completion time: **MTTTIME**
◦ Correct lines: **MTCORR**
◦ Errors: **MTERROR**
3. **Digit Span:**

◦ Maximum forward recall span: **DIGITFW**
◦ Maximum backward recall span: **DIGITBW**
4. **Peabody Picture Vocabulary Test (PPVT):**

◦ Verbal category: **PPVTVRB**
◦ Descriptive category: **PPVTDES**
◦ Animate category: **PPVTANI**
◦ Inanimate category: **PPVTINA**

This study includes the BNT, Modified Trails, Digit Span, and PPVT to evaluate cognitive domains commonly affected in FTD and its subtypes. The BNT assesses naming ability for visually presented objects, with lower scores strongly linked to semantic deficits and anterior temporal lobe atrophy, particularly in svFTD [29]. Modified Trails evaluates executive functioning, with errors and slower completion times reflecting impaired attention and cognitive flexibility, key deficits in bvFTD [30] . Digit Span measures working memory, with backward recall highlighting executive dysfunction associated with early frontal atrophy. The PPVT captures semantic deficits across verbal, descriptive, animate, and inanimate categories, indicative of anterior temporal lobe degeneration in svFTD [29] . Partial correlations between thalamic nuclei volumes and cognitive performance (measured by BNTCORR, MTTTIME, MTCORR, MTERROR, DIGITFW, DIGITBW, PPVTVRB, PPVTDES, PPVTANI, and PPVTINA scores) were examined while controlling for age, sex, and ICV to uncover disease-specific patterns and explore the neural mechanisms underlying FTD symptoms.

## 3. RESULTS

### 3.1. Descriptive Statistics

At Timepoint 1 (baseline), the study included 274 subjects, comprising 145 males (52.9%) and 129 females (47.1%). The group distribution was as follows: 65 bvFTD cases (23.7%), 136 controls (49.6%), 36 PNFA cases (13.1%), and 37 svFTD cases (13.5%). The overall mean age at baseline was 63.2 ± 7.28 years (range: 36–81 years). Group-wise, the mean age for bvFTD was 61.2 ± 6.55 years (45–74 years), controls were 63.0 ± 7.38 years (36–81 years), PNFA cases were 68.0 ± 7.33 years (54–81 years), and svFTD cases were 63.1 ± 6.16 years (50–73 years).

At Timepoint 2 (follow-up), 237 subjects remained in the study, including 124 males (52.3%) and 113 females (47.7%). The group distribution was 50 bvFTD cases (21.1%), 122 controls (51.5%), 31 PNFA cases (13.1%), and 34 svFTD cases (14.3%). The overall mean age at follow-up was 65.8 ± 7.79 years (range: 37–85 years). Within groups, the mean age for bvFTD was 62.7 ± 6.48 years (48–76 years), controls were 66.5 ± 8.17 years (37–85 years), PNFA cases were 69.2 ± 7.96 years (55–83 years), and svFTD cases were 65 ± 6.37 years (52–75 years) **(Table 2)**.

**Table 2.** Summary of demographic and age-related characteristics of participants at timepoint 1 (baseline) and timepoint 2 (longitudinal follow-up). It includes total subjects, gender distribution, group composition (bvFTD, CN, PNFA, svFTD), and overall and group-specific age statistics (mean ± SD, range).

| Metric | Timepoint 1 | Timepoint 2 |
| --- | --- | --- |
| Total Subjects | 274 | 237 |
| Males (Count, %) | 145 (52.9%) | 124 (52.3%) |
| Females (Count, %) | 129 (47.1%) | 113 (47.7%) |
| BV (% of total) | 65 (23.7%) | 50 (21.1%) |
| CN (% of total) | 136 (49.6%) | 122 (51.5%) |
| PNFA (% of total) | 36 (13.1%) | 31 (13.1%) |
| SV (% of total) | 37 (13.5%) | 34 (14.3%) |
| Age (Overall) (Mean ± SD, Range) | 63.2 ± 7.28 (36-81) | 65.8 ± 7.79 (37-85) |
| Age by Group (CN) (Mean ± SD, Range) | 63.0 ± 7.38 (36-81) | 66.5 ± 8.17 (37-85) |
| Age by Group (bvFTD) (Mean ± SD, Range) | 61.2 ± 6.55 (45-74) | 62.7 ± 6.48 (48-76) |
| Age by Group (PNFA) (Mean ± SD, Range) | 68.0 ± 7.33 (54-81) | 69.2 ± 7.96 (55-83) |
| Age by Group (svFTD) (Mean ± SD, Range) | 63.1 ± 6.16 (50-73) | 65 ± 6.37 (52-75) |

The 274 subjects at Timepoint 1 and 237 subjects at Timepoint 2 included only those who passed quality control (QC). Initially, 281 subjects were available at Timepoint 1, but six subjects failed QC due to segmentation failure on either the left or right side, or both. At Timepoint 2, 244 subjects were initially available, but 7 subjects failed QC for similar reasons. Additionally, within the final datasets of 274 and 237 subjects, some cases had basal ganglia segmentations that failed specifically in the caudate and nucleus accumbens; these values were blanked out. Since outlier detection was performed separately for each nucleus, any such cases were accounted for accordingly.

### 3.2. Thalamic Volume Atrophy

#### 3.2.1. Cross-sectional Analysis across FTD subtypes

Our cross-sectional analysis at Timepoint 1 (baseline) revealed significant structural differences in specific thalamic and deep gray matter nuclei across FTD subtypes. The MD nucleus consistently exhibited the strongest effect sizes, particularly in bvFTD, with Cohen’s d = 2.043 (p < 0.001) in the left hemisphere and 2.282 (p < 0.001) in the right hemisphere, highlighting substantial volume loss. The AV nucleus also showed notable reductions in bvFTD (left: d = 1.84, right: d = 1.718, p < 0.001), suggesting early involvement in this subtype (**Figure 2 and Table 3**).

**Figure 2.**
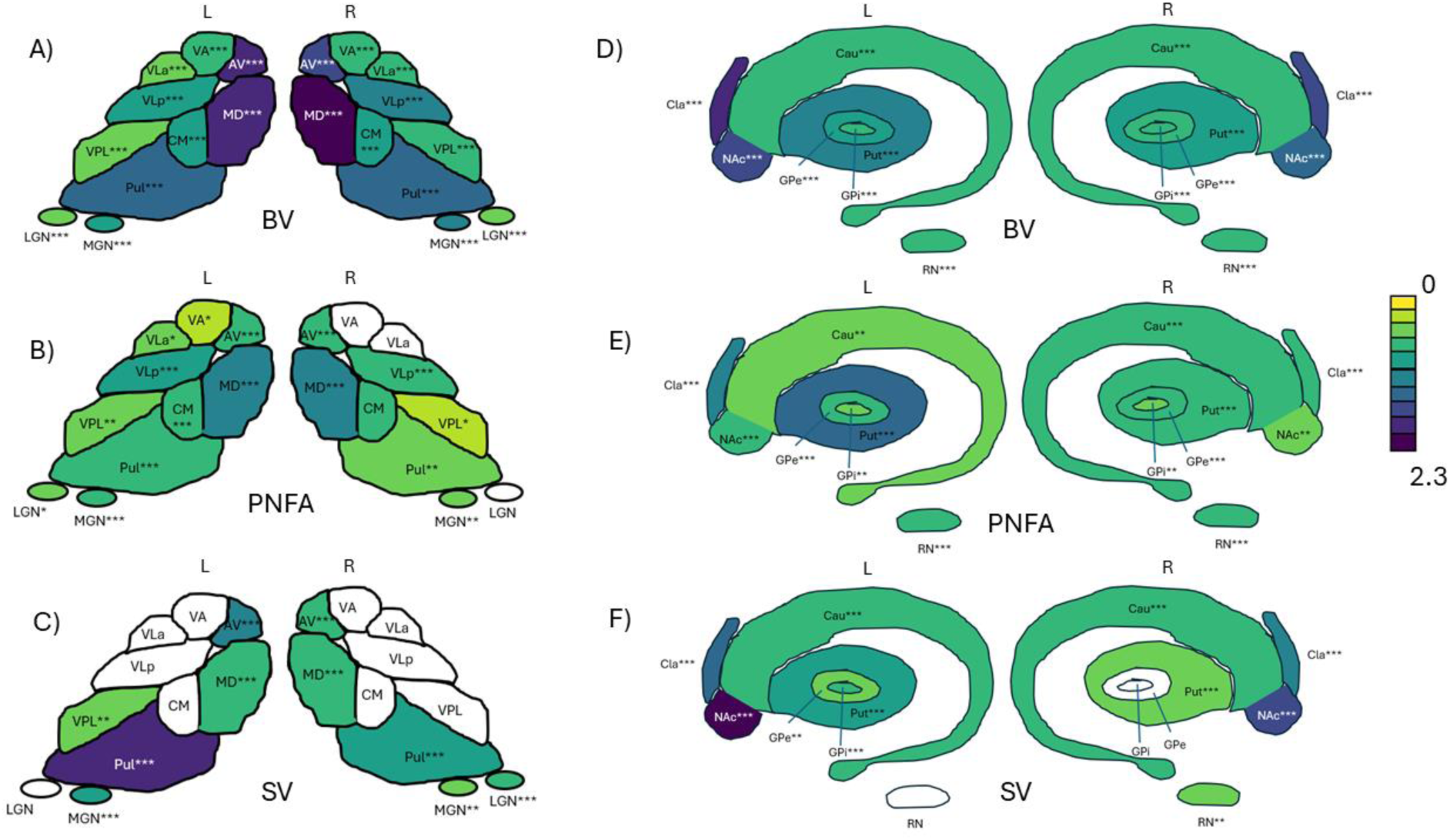
Schematic showing significant effect sizes (Cohen’s d) in thalamic and deep grey regions for timepoint 1 (baseline) Panels (A) and (D) represent bvFTD vs CN, demonstrating the most pronounced effect sizes in regions like MD (Cohen’s d = 2.043, p < 0.001) and AV (Cohen’s d = 1.84, p < 0.001) in the left hemisphere. Similarly, in the right hemisphere, MD exhibited the highest effect size (Cohen’s d = 2.282, p < 0.001). Panels (B) and (E) illustrate PNFA vs CN, showing notable alterations in the left putamen (Cohen’s d = 1.387, p < 0.001). Panels (C) and (F) represent svFTD vs CN, with substantial differences observed in the Pul (Cohen’s d = 1.692, p < 0.001) and MD. The accumbens and caudate consistently showed moderate to high effect sizes, emphasizing their relevance across FTD subtypes.

The Pul nucleus displayed significant atrophy in both bvFTD (left: d = 1.487, right: d = 1.563, p < 0.001) and svFTD (left: d = 1.692, right: d = 1.105, p < 0.001), suggesting shared vulnerabilities. The putamen and nucleus accumbens were prominently affected in bvFTD and PNFA, while the caudate exhibited moderate atrophy (bvFTD, left: d = 0.847, right: d = 0.781, p < 0.001). These findings suggest that bvFTD shows the most widespread involvement, svFTD has distinct pulvinar and accumbens reductions, and PNFA exhibits more selective involvement of the putamen and caudate (**Figure 2 and Table 3 and 4**).

**Table 3.**
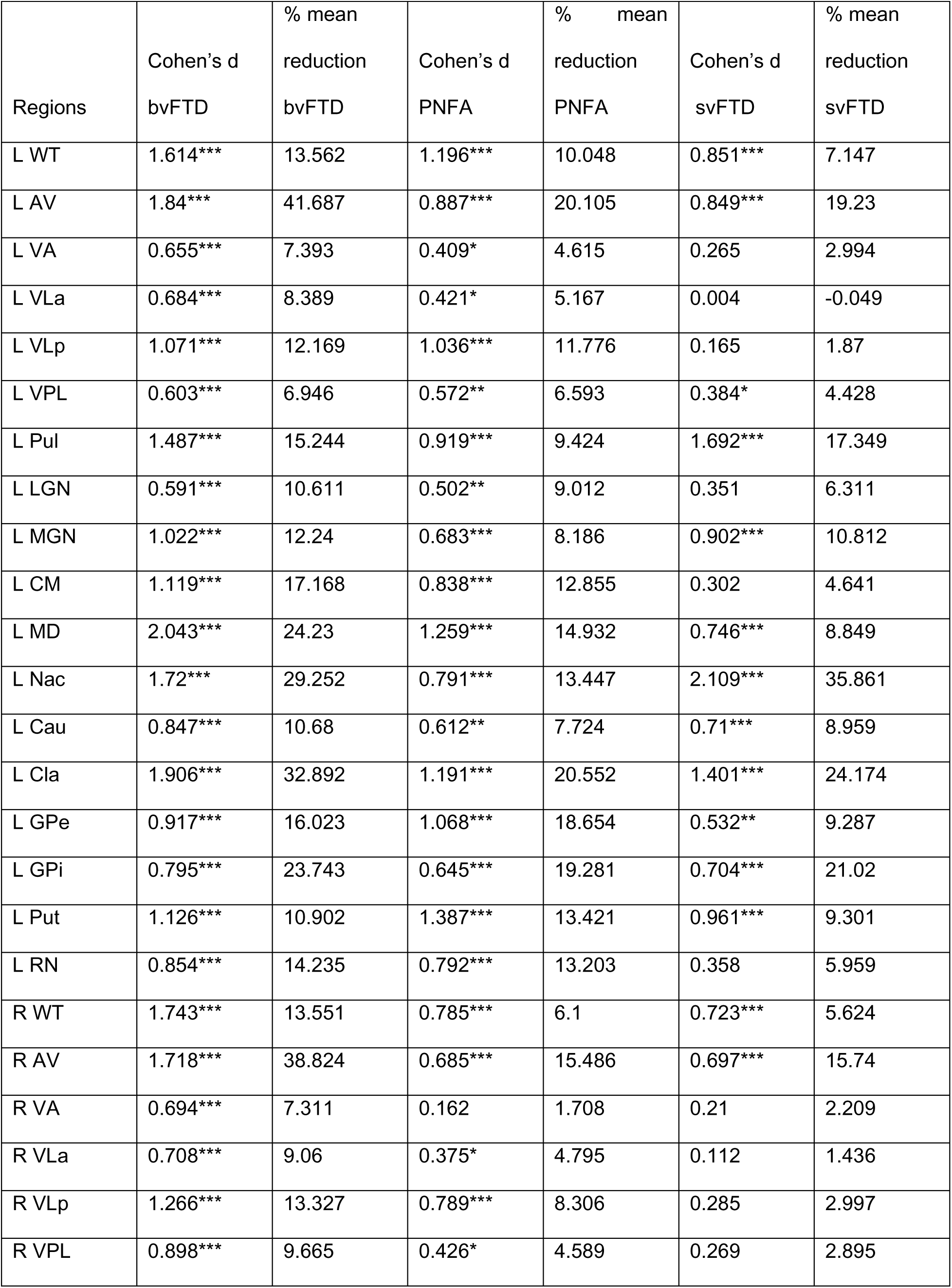
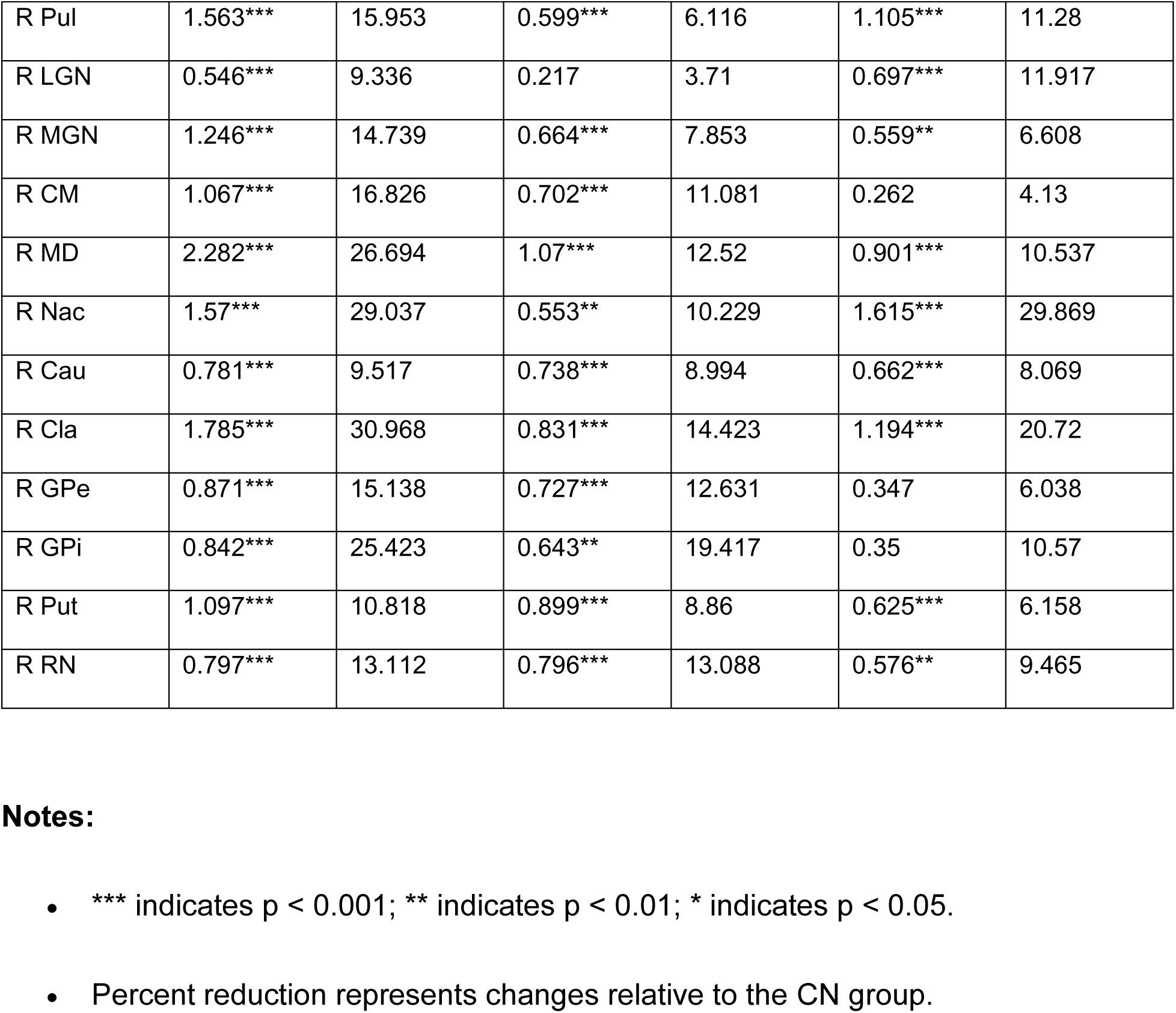
Comparison of Cohen’s d Across Groups at Timepoint 1 (Baseline) Cohen’s d effect sizes and percent volume reductions comparing structural changes in thalamic and deep gray matter nuclei among three clinical groups (bvFTD, PNFA, and svFTD) relative to the control group (CN) at timepoint 1 (baseline). The table provides detailed comparisons across key thalamic and deep gray matter regions in both hemispheres, quantifying the extent of atrophy and its progression over time. Effect sizes (Cohen’s d) indicate the magnitude of structural differences, with statistical significance denoted by asterisks: *** (p < 0.001), ** (p < 0.01), and * (p < 0.05). Percent reduction values reflect the relative decline in mean volume for each clinical group compared to CN at each timepoint.

| Regions | Cohen's d<br>bvFTD | % mean<br>reduction<br>bvFTD | Cohen's d<br>PNFA | % mean<br>reduction<br>PNFA | Cohen's d<br>svFTD | % mean<br>reduction<br>svFTD |
| --- | --- | --- | --- | --- | --- | --- |
| L WT | 1.614*** | 13.562 | 1.196*** | 10.048 | 0.851*** | 7.147 |
| L AV | 1.84*** | 41.687 | 0.887*** | 20.105 | 0.849*** | 19.23 |
| L VA | 0.655*** | 7.393 | 0.409* | 4.615 | 0.265 | 2.994 |
| L VLa | 0.684*** | 8.389 | 0.421* | 5.167 | 0.004 | -0.049 |
| L VLp | 1.071*** | 12.169 | 1.036*** | 11.776 | 0.165 | 1.87 |
| L VPL | 0.603*** | 6.946 | 0.572** | 6.593 | 0.384* | 4.428 |
| L Pul | 1.487*** | 15.244 | 0.919*** | 9.424 | 1.692*** | 17.349 |
| L LGN | 0.591*** | 10.611 | 0.502** | 9.012 | 0.351 | 6.311 |
| L MGN | 1.022*** | 12.24 | 0.683*** | 8.186 | 0.902*** | 10.812 |
| L CM | 1.119*** | 17.168 | 0.838*** | 12.855 | 0.302 | 4.641 |
| L MD | 2.043*** | 24.23 | 1.259*** | 14.932 | 0.746*** | 8.849 |
| L Nac | 1.72*** | 29.252 | 0.791*** | 13.447 | 2.109*** | 35.861 |
| L Cau | 0.847*** | 10.68 | 0.612** | 7.724 | 0.71*** | 8.959 |
| L Cla | 1.906*** | 32.892 | 1.191*** | 20.552 | 1.401*** | 24.174 |
| L GPe | 0.917*** | 16.023 | 1.068*** | 18.654 | 0.532** | 9.287 |
| L GPi | 0.795*** | 23.743 | 0.645*** | 19.281 | 0.704*** | 21.02 |
| L Put | 1.126*** | 10.902 | 1.387*** | 13.421 | 0.961*** | 9.301 |
| L RN | 0.854*** | 14.235 | 0.792*** | 13.203 | 0.358 | 5.959 |
| R WT | 1.743*** | 13.551 | 0.785*** | 6.1 | 0.723*** | 5.624 |
| R AV | 1.718*** | 38.824 | 0.685*** | 15.486 | 0.697*** | 15.74 |
| R VA | 0.694*** | 7.311 | 0.162 | 1.708 | 0.21 | 2.209 |
| R VLa | 0.708*** | 9.06 | 0.375* | 4.795 | 0.112 | 1.436 |
| R VLp | 1.266*** | 13.327 | 0.789*** | 8.306 | 0.285 | 2.997 |
| R VPL | 0.898*** | 9.665 | 0.426* | 4.589 | 0.269 | 2.895 |
| R Pul | 1.563*** | 15.953 | 0.599*** | 6.116 | 1.105*** | 11.28 |
| R LGN | 0.546*** | 9.336 | 0.217 | 3.71 | 0.697*** | 11.917 |
| R MGN | 1.246*** | 14.739 | 0.664*** | 7.853 | 0.559** | 6.608 |
| R CM | 1.067*** | 16.826 | 0.702*** | 11.081 | 0.262 | 4.13 |
| R MD | 2.282*** | 26.694 | 1.07*** | 12.52 | 0.901*** | 10.537 |
| R Nac | 1.57*** | 29.037 | 0.553** | 10.229 | 1.615*** | 29.869 |
| R Cau | 0.781*** | 9.517 | 0.738*** | 8.994 | 0.662*** | 8.069 |
| R Cla | 1.785*** | 30.968 | 0.831*** | 14.423 | 1.194*** | 20.72 |
| R GPe | 0.871*** | 15.138 | 0.727*** | 12.631 | 0.347 | 6.038 |
| R GPi | 0.842*** | 25.423 | 0.643** | 19.417 | 0.35 | 10.57 |
| R Put | 1.097*** | 10.818 | 0.899*** | 8.86 | 0.625*** | 6.158 |
| R RN | 0.797*** | 13.112 | 0.796*** | 13.088 | 0.576** | 9.465 |

**Table 4.**
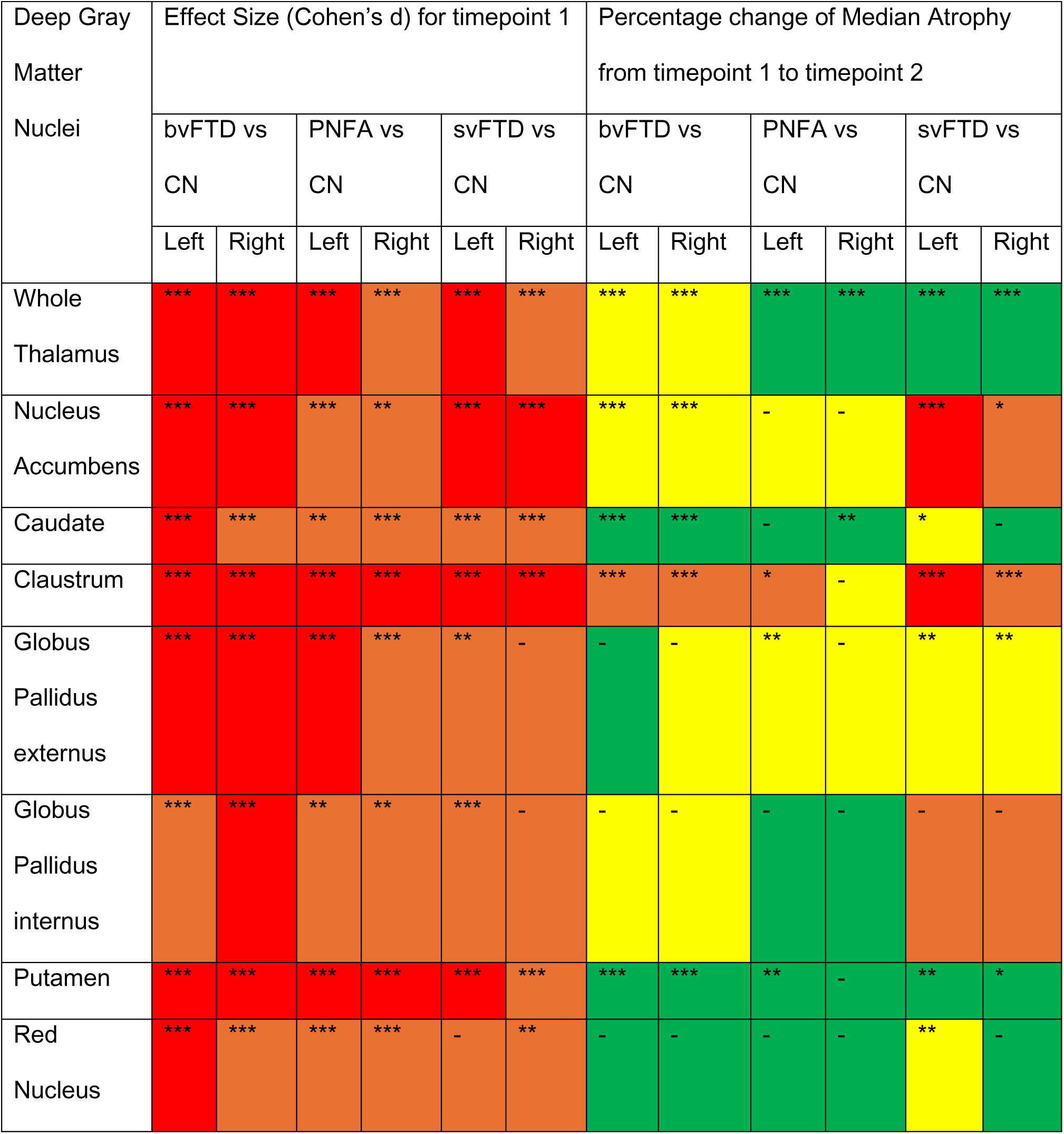
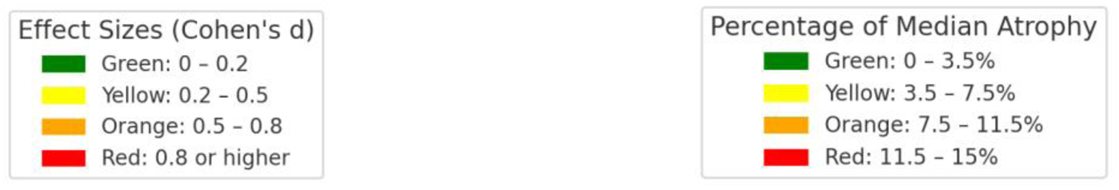
Comparison of effect sizes (Cohen’s d) of deep gray matter nuclei structure volume differences and the percentage change in median atrophy between bvFTD, PNFA, svFTD, and CN groups. Effect sizes are color-coded: green (0–0.2), yellow (0.2– 0.5), orange (0.5–0.8), and red (≥0.8). Median atrophy percentage changes are similarly color-coded: green (0–3.5%), yellow (3.5–7.5%), orange (7.5–11.5%), and red (11.5– 15%). Statistical significance is denoted by *** (p < 0.001), ** (p < 0.01), and * (p < 0.05).

Across both timepoints, the WT shows the highest percentage reductions of adjusted means, particularly in bvFTD (TP1: L 13.56%, R 13.55%; TP2: L 17.98%, R 16.93%) and PNFA (TP1: L 10.05%, R 6.10%; TP2: L 14.14%, R 8.62%). Among thalamic nuclei, the AV nucleus exhibits the most severe atrophy, with bvFTD (TP1: L 41.69%, R 38.82%; TP2: L 55.91%, R 48.45%) and svFTD (TP1: L 19.23%, R 15.74%; TP2: L 32.52%, R 29.78%), while the MD is also highly affected, particularly in bvFTD (TP1: L 24.23%, R 26.69%; TP2: L 32.02%, R 29.62%) and PNFA (TP1: L 14.93%, R 12.52%; TP2: L 24.10%, R 16.52%). In other deep gray matter nuclei, the NAc shows severe atrophy in svFTD (TP1: L 35.86%, R 29.87%; TP2: L 47.90%, R 42.33%) and bvFTD (TP1: L 29.25%, R 29.04%; TP2: L 34.16%, R 33.13%), while the Cla also exhibits significant degeneration, with bvFTD (TP1: L 32.89%, R 30.97%; TP2: L 40.19%, R 39.64%) and svFTD (TP1: L 24.17%, R 20.72%; TP2: L 39.68%, R 31.21%), highlighting cortical integration deficits (**Table 3 and Supplementary Table 2**).

#### 3.2.2. Longitudinal Analysis of Atrophy

Our results demonstrate significant and pronounced atrophy across multiple thalamic and deep gray matter nuclei in disease groups compared to controls, with distinct patterns of regional atrophy for each subtype (**Figure 3**).

**Figure 3.**
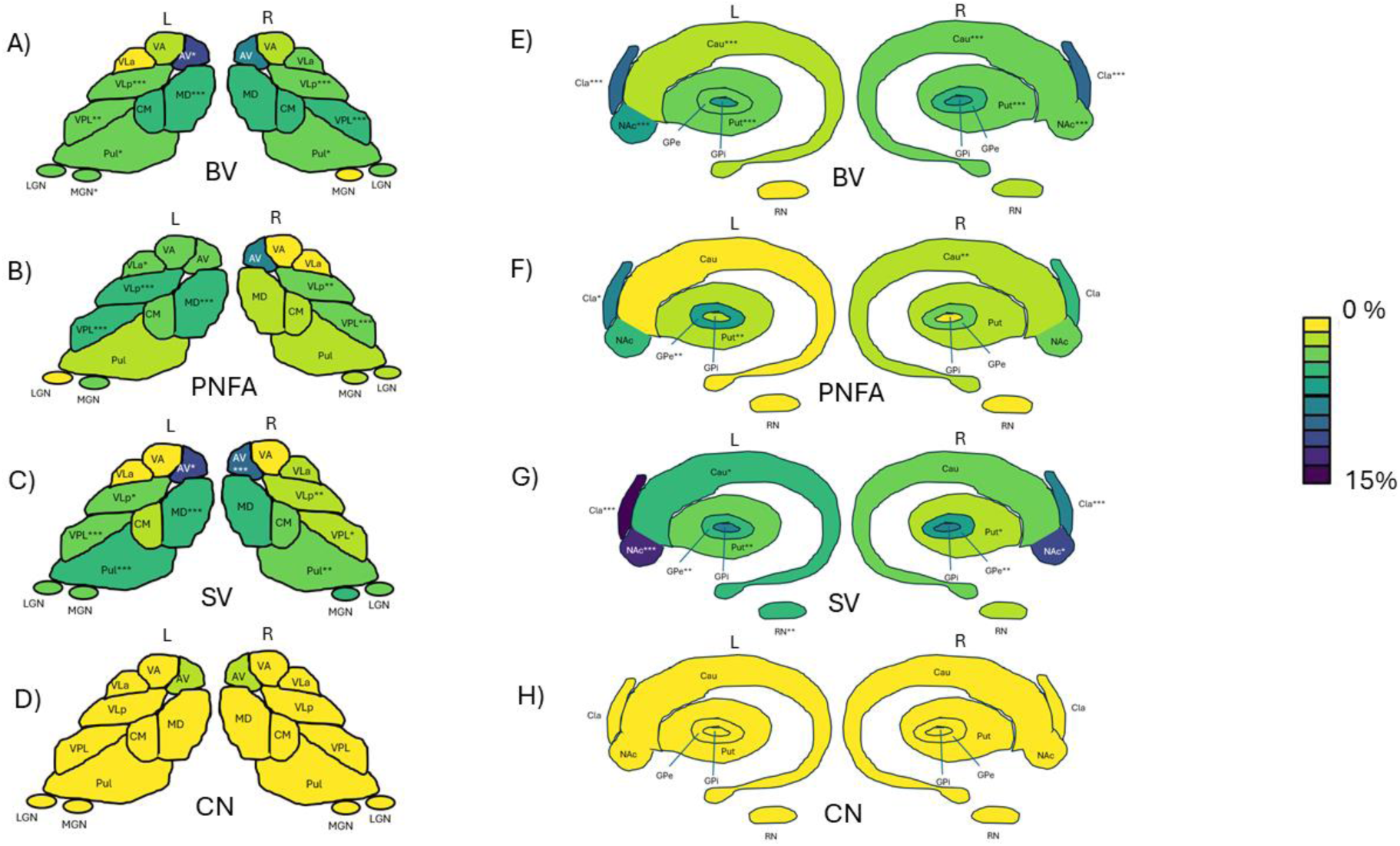
Group-Wise Differences in Median Atrophy Across Thalamic and other Deep Grey Matter Regions. Percentage of median atrophy and statistical significance across thalamic and deep gray matter nuclei for bvFTD vs CN (A, E), PNFA vs CN (B, F), and svFTD vs CN (C, G) and CN vs CN (D, H). The analysis reveals distinct patterns of atrophy, with bvFTD showing the most significant and widespread differences, particularly in the AV, MD, and Pul nuclei of the thalamus, as well as other deep gray matter nuclei such as the nucleus accumbens and putamen. The pulvinar nucleus exhibited consistent vulnerability across all FTD subtypes, highlighting its importance in disease progression. Statistical annotations (*p < 0.05, **p < 0.01, ***p < 0.001) indicate significant differences compared to the control group. The color bar represents the range of median atrophy, spanning 0–15%.

In bvFTD, significant bilateral atrophy was observed in the whole thalamus, with greater right hemisphere reductions (4.08%, p < 0.00001) compared to the left (3.68%, p < 0.00001). The MD nucleus (left: 5.37%, p < 0.001) and VLp (left: 3.82%, p < 0.0001) showed substantial atrophy, while the nucleus accumbens (left: 6.64%, p < 0.0001) and claustrum (left: 10.3%, p < 0.0001) exhibited the most pronounced reductions, indicating progressive degeneration beyond baseline differences.

In PNFA, atrophy was more localized, primarily affecting the left VLp (5.26%, p < 0.0001) and left MD (5.62%, p < 0.001), with additional reductions in the right caudate (2.47%, p < 0.01) and right putamen (2.44%, p < 0.01), suggesting a slower and more regionally restricted progression.

In svFTD, bilateral atrophy was observed, particularly in the whole thalamus (left: 2.72%, p < 0.00001; right: 4.08%, p < 0.00001) and nucleus accumbens (left: 12.9%, p < 0.001; right: 10.5%, p < 0.05). Significant reductions were also noted in the left AV (11.3%, p < 0.01), left MD (4.87%, p < 0.001), left Pul (5.19%, p < 0.001), and left claustrum (14.4%, p < 0.0001).

Cross-sectional analysis in this study highlights early structural vulnerabilities, particularly in the MD, AV, Pul, and nucleus accumbens, where bvFTD shows the most pronounced baseline differences. However, longitudinal analysis reveals progressive degeneration that was not evident cross-sectionally, such as substantial atrophy in the claustrum and caudate, which had only moderate baseline differences. Additionally, while the putamen and accumbens appear highly affected at baseline in bvFTD, their longitudinal decline is less steep, suggesting early but more stable degeneration. In contrast, the AV nucleus, which showed significant cross-sectional reductions in bvFTD, exhibits progressive atrophy in svFTD, indicating subtype-specific disease trajectories. These findings underscore the importance of cross-sectional and longitudinal data for identifying early disease markers and longitudinal data for tracking progression and differentiating stable from degenerating regions.

### 3.3. Partial correlation analyses

Significant positive partial correlations were observed between thalamic and other deep gray matter structures, and neuropsychological measures in different subtypes of FTD. In bvFTD, the ventral lateral posterior (VLp) nucleus in both hemispheres demonstrated positive correlations with working memory performance, as assessed by DIGITBW (left: 𝑟 = 0.307, 𝑝 = 0.0161; right: 𝑟 = 0.284, 𝑝 = 0.0268). The accumbens was significantly associated with naming ability, measured by BNTCORR, with positive correlations in the left (𝑟 = 0.335, 𝑝 = 0.0102) and right hemispheres (𝑟 = 0.340, 𝑝 = 0.0090). The claustrum showed positive correlations with semantic task performance (PPVTANI) in both hemispheres (left: 𝑟 = 0.372, 𝑝 = 0.0140; right: 𝑟 = 0.313, 𝑝 = 0.0407). Additionally, the right mediodorsal (MD) nucleus correlated significantly with naming ability (BNTCORR: 𝑟 = 0.336, 𝑝 = 0.0081) (**Table 5**).

**Table 5.**
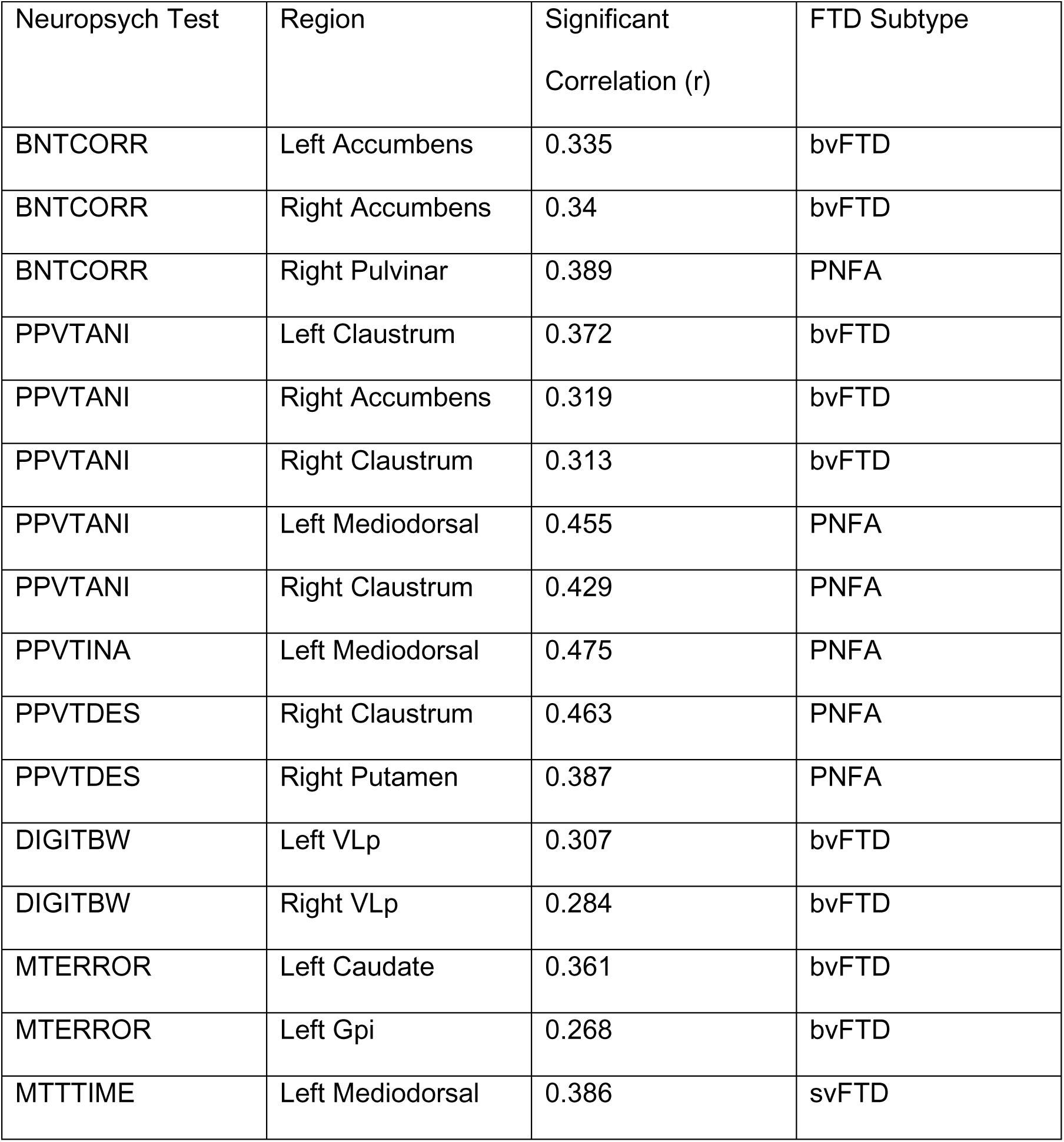
Significant Positive Correlations Between Neuropsychological Tests and Brain Regions in FTD Subtypes. Highlights significant positive correlations observed between specific neuropsychological tests and thalamic or deep gray matter nuclei across three FTD subtypes: bvFTD, PNFA, and svFTD. The results are categorized by neuropsychological tests, including BNTCORR, PPVTANI, PPVTINA, PPVTDES, DIGITBW, MTERROR, and MTTTIME. Each correlation is reported with its strength (pearson’s r-value) and significance level (* for p < 0.05, ** for p < 0.01, *** for p < 0.001). This analysis reveals the involvement of distinct thalamic and deep gray matter structures in mediating cognitive and behavioral functions specific to each FTD subtype.

In PNFA, the left mediodorsal nucleus exhibited strong positive correlations with semantic tasks, including PPVTANI (𝑟 = 0.455, 𝑝 = 0.0171) and PPVTINA (𝑟 = 0.475, 𝑝 = 0.0123). The right pulvinar nucleus was significantly associated with naming performance (BNTCORR: 𝑟 = 0.629, 𝑝 = 0.0001), visuospatial processing (MTTTIME: 𝑟 = 0.444, 𝑝 = 0.0108), and task accuracy (MTERROR: 𝑟 = 0.416, 𝑝 = 0.0177). The right claustrum demonstrated significant correlations with descriptive and semantic tasks, including PPVTDES (𝑟 = 0.463, 𝑝 = 0.0130) and PPVTANI (𝑟 = 0.429, 𝑝 = 0.0228).

In svFTD, the left mediodorsal nucleus showed a positive correlation with task completion time (MTTTIME: 𝑟 = 0.386, 𝑝 = 0.0350). These findings highlight distinct patterns of thalamic and other deep gray matter l nuclei involvement in cognitive and neuropsychological deficits across bvFTD, PNFA, and svFTD subtypes, underscoring their role in disease-specific cognitive impairments.

Moreover, all partial correlation results, including significant values, are presented in **Table 5**, while Pearson correlation results are compiled in **Supplementary Table 3**, which summarizes the Pearson correlations, highlighting notable structural-functional relationships. For instance, in bvFTD, significant correlations were observed between DIGITBW and the right VLp, as well as between BNTCORR and the left mediodorsal nucleus. In PNFA, strong correlations were found for PPVTVRB with the left mediodorsal nucleus and left claustrum, and for PPVTANI with the left pulvinar and right accumbens. Notably, the highest correlation was seen for PPVTINA with the left mediodorsal nucleus in PNFA. These results collectively offer insights into disease-specific structural-functional patterns and underscore the role of deep gray matter nuclei in cognitive and behavioral impairments.

### 3.4. Sex specific patterns

Although our primary focus was on group-level differences, we also examined sex-specific patterns in atrophy across FTD subtypes. Thus, additionally, analyses were stratified by sex to examine male-female differences in atrophy trajectories, ensuring that distinct neurodegenerative patterns between sexes were captured in each clinical group, with the analysis conducted in the longitudinal framework to account for changes over time and assess sex-specific atrophy trajectories.

Cross-sectionally, females exhibited greater atrophy in right AV (d = 2.377, p < 0.001) and nucleus accumbens (d = 2.385, p < 0.001) in bvFTD, while males showed more pronounced reductions in the left AV (d = 1.725, p < 0.001) and left MD (d = 1.874, p < 0.001) in timepoint 1. In svFTD, females had more severe pulvinar atrophy (d = 2.133, p < 0.001) compared to males (d = 1.424, p < 0.001), whereas PNFA showed fewer sex differences, with both sexes exhibiting comparable putamen atrophy.

Longitudinally, females exhibited more progressive atrophy in the nucleus accumbens, with severe reductions in left NAc (bvFTD: 14.1%, p < 0.001; svFTD: 15.1%, p < 0.01) and right NAc (bvFTD: 10.5%, p < 0.001), whereas males showed only mild effects (left NAc bvFTD: 4.94%, p < 0.01). Conversely, males exhibited stronger longitudinal atrophy in the caudate (right Caudate bvFTD: 2.58%, p < 0.001; PNFA: 3.12%, p < 0.05) and putamen (left Putamen bvFTD: 3.34%), while females showed only svFTD-related caudate reductions (left Caudate: 4.57%, p < 0.05). The claustrum displayed bilateral atrophy in females but was more asymmetric in males, primarily affecting left Claustrum (bvFTD: 10.1%; svFTD: 13.5%, p < 0.001).

These findings suggest sex-dependent vulnerabilities, with females exhibiting more severe atrophy in some deep gray matter nuclei, while males show more pronounced caudate and putamen degeneration. However, we do not have a strong hypothesis for these sex differences, and given the sample size limitations, these results should be interpreted with caution. Future studies with larger cohorts and meta-analyses will be necessary to better characterize sex-specific neurodegenerative trajectories in FTD. Full results are available in the Supplemental Materials (**Supplementary figure 2 - 5**)

## 4. DISCUSSION

This study introduces the novel use of the sTHOMAS segmentation method to analyze deep gray matter nuclei—including the thalamus, basal ganglia, claustrum, and red nuclei—in FTD. By synthesizing WMn-like contrast from standard T1-weighted MRI, sTHOMAS overcomes the contrast limitations of traditional T1 contrast. This is the first study to incorporate both cross-sectional and longitudinal analyses of atrophy in FTD subtypes, with an additional examination of gender differences. Our findings highlight the progressive degeneration of deep gray matter, with significant atrophy in the MD, AV, pulvinar, and VLp thalamic nuclei, as well as in the nucleus accumbens, caudate, and putamen, particularly in bvFTD and svFTD.

Consistent with previous studies [7] [12] [17] [42], our results confirm the widespread involvement of thalamic nuclei in bvFTD and the selective atrophy patterns in PNFA and svFTD. Notably, significant volume reductions were observed in the MD, Pulvinar, and AV nuclei across all patient groups, with the most pronounced atrophy in bvFTD. These findings corroborate the work of [12], which identified MD nucleus involvement as a hallmark feature across FTD subtypes. The MD nucleus, central to executive, emotional, and behavioral regulation through its connections with prefrontal, temporal, and limbic regions, was universally affected across our cohort, underscoring its pivotal role in FTD pathophysiology.

McKenna et al. (2022) provided a comprehensive characterization of thalamic atrophy in FTD subtypes, highlighting phenotype-specific focal degeneration and thalamocortical circuit disruption. Similarly, Bocchetta et al. (2020) used FreeSurfer for thalamic nuclei segmentation. However, both studies had limitations.. While McKenna et al. included 170 participants, their FTD cohort was relatively small (n = 70) and unevenly distributed across subtypes, with particularly low representation of svPPA (n = 5), bvFTD (n = 10), and nfvPPA (n = 15). This imbalance limits the statistical power for subtype-specific analyses. Moreover, neither study incorporated longitudinal imaging, focusing solely on cross-sectional data. This lack of temporal analysis restricts insights into the progression of thalamic degeneration over time. In contrast, our study integrates both cross-sectional and longitudinal analyses, providing a more comprehensive view of disease progression. By including median atrophy rate assessments, we capture the progressive nature of thalamic degeneration across FTD subtypes.

Our findings diverge significantly from those of Bocchetta et al. (2020), who examined thalamic nuclei involvement in FTD subtypes using a larger cohort (n = 402). While their study identified widespread MD nucleus involvement across FTD subtypes and highlighted the selective vulnerability of the pulvinar in C9orf72-associated FTD, it did not investigate longitudinal changes or sex-specific patterns of degeneration. In contrast, our study reveals that the pulvinar and VLp nuclei are particularly affected in svFTD and PNFA, with more pronounced atrophy in males compared to females. By incorporating longitudinal median atrophy analysis, we provide new insights into the dynamic nature of thalamic degeneration, offering a more comprehensive understanding of disease progression.

Our longitudinal analysis reveals a gradient of volume reduction in FTD, with bvFTD showing the most extensive bilateral atrophy, svFTD exhibiting targeted reductions, and PNFA displaying more focal and asymmetric patterns. A distinct left-lateralized atrophy pattern emerges in key thalamic and other deep gray nuclei regions, including the AV, MD, and VLp nuclei, caudate, claustrum, and nucleus accumbens, likely reflecting the left hemisphere’s role in language, memory, and emotional regulation. In bvFTD, widespread atrophy in the MD, VLp, and pulvinar nuclei, along with the nucleus accumbens, caudate, and claustrum, aligns with frontostriatal and limbic circuit disruptions [31]. The MD nucleus’s prefrontal connections implicate executive dysfunction [33], while pulvinar involvement suggests visuospatial and attentional deficits [34]. In PNFA, left-lateralized atrophy in the VLp, MD, putamen, and GPe corresponds with language deficits, affecting motor planning and speech production [35]. In svFTD, bilateral AV, Pulvinar, and MD atrophy disrupts the semantic memory network and limbic structures [36].

Our findings reveal sex-specific neuroanatomical vulnerabilities in FTD, with females showing greater nucleus accumbens and claustrum atrophy, particularly in bvFTD and svFTD. Given their roles in reward processing and sensory integration, this may contribute to greater apathy and attentional deficits in females. In contrast, males exhibited more caudate and putamen atrophy, particularly in bvFTD and PNFA, suggesting greater executive dysfunction and motor rigidity. Thalamic nuclei findings further highlight sex differences, with females showing more AV atrophy in PNFA and svFTD, linked to worse memory and language deficits, while males had greater MD atrophy in bvFTD, associated with executive dysfunction [27]. The VLp and pulvinar nuclei showed subtype-specific patterns, with males exhibiting more VLp atrophy in PNFA and females showing greater pulvinar degeneration in bvFTD. However, the underlying mechanisms driving these differences remain unclear. Given the sample size limitations, these results should be interpreted with caution. Future studies with larger cohorts and meta-analyses will be essential to further characterize sex-specific vulnerabilities and their impact on clinical presentation in FTD.Our study uniquely integrates both structural and functional analyses to examine the cognitive and behavioral implications of subcortical degeneration in FTD. While McKenna et al. (2022) and Bocchetta et al. (2020) focused primarily on structural changes, our partial correlation analysis links thalamic nuclei volumes to neuropsychological performance across FTD subtypes. We found that the VLp nucleus positively correlated with working memory performance (DIGITBW) in bvFTD, while the MD nucleus was associated with semantic and naming tasks (BNTCORR, PPVTANI) in PNFA. The nucleus accumbens showed significant positive correlations with naming ability (BNTCORR) in bvFTD and with semantic processing (PPVTANI) in both bvFTD and PNFA. The claustrum was also linked to semantic tasks (PPVTANI, PPVTDES), reinforcing its role in language processing. Additionally, basal ganglia structures, including the caudate and putamen, were associated with visuospatial task accuracy (MTERROR) in bvFTD.

Previous studies [37] [38] have highlighted that cross-sectional analyses often underestimate true brain changes, a limitation our findings confirm. The discrepancies between cross-sectional and longitudinal data reinforce the necessity of capturing temporal dynamics to accurately map neurodegenerative trajectories. Our results particularly emphasize the rapid degeneration in bvFTD, with the nucleus accumbens and pulvinar showing continuous decline over time.

The claustrum, often described as one of the brain’s most interconnected structures, receives and projects information to nearly all cortical regions, supporting its proposed role in integrating multimodal sensory, motor, and cognitive processes [39]. [40] hypothesized that the claustrum functions as a "conductor of consciousness," unifying perceptual experience through its extensive connectivity. Given this role, its degeneration in FTD may contribute to the disorder’s diverse cognitive and behavioral deficits by disrupting these integrative processes. Our findings underscore the need for future studies to explore the functional consequences of claustral atrophy in FTD, particularly in relation to attention, awareness, and cognitive integration.

Moreover, the selective nature of thalamic degeneration in FTD was evident in both cross-sectional and longitudinal analyses. While the AV, MD, and pulvinar nuclei were significantly affected, other regions such as the CM, LGN, and MGN showed non-significant changes, underscoring the heterogeneous vulnerability of thalamic subregions.

While our study provides valuable insights into the selective and dynamic atrophy of thalamic nuclei in FTD, a few limitations should be acknowledged. First, the smaller sample sizes for PNFA (n = 36) and svFTD (n = 37) compared to bvFTD may reduce statistical power, potentially biasing results toward more pronounced atrophy patterns in bvFTD. This disparity may also contribute to differences between cross-sectional and longitudinal findings, emphasizing the need for larger, balanced cohorts across FTD subtypes to improve statistical robustness and generalizability.

Additionally, while the sTHOMAS segmentation method improves accuracy, the NIFD dataset was collected from three sites with slightly varied imaging protocols. Although most data were acquired from a single scanner, minimizing site effects, future studies using multi-site data should incorporate harmonization techniques to account for differences in scanner models, acquisition parameters, and voxel sizes, which could otherwise introduce variability and impact segmentation accuracy.

Moreover, our study focused exclusively on volumetric atrophy, without integrating complementary modalities such as diffusion-weighted imaging, which could offer deeper insights into microstructural changes and connectivity disruptions in thalamocortical circuits. Future research should incorporate multi-modal imaging approaches, including DTI and functional MRI, to better understand the structural and functional consequences of thalamic degeneration in FTD. Additionally, while we identified sex-specific differences, these findings should be interpreted with caution given the exploratory nature of the analysis.

In conclusion, our study provides novel insights into the selective and progressive atrophy of deep gray matter structures in FTD, highlighting the involvement of thalamic nuclei, regions of the basal ganglia, and the claustrum. By integrating both cross-sectional and longitudinal analyses, we reveal distinct subtype- and sex-specific patterns of degeneration, underscoring the dynamic nature of thalamic and basal ganglia involvement in FTD progression. The strong correlations between structural atrophy and neuropsychological performance further emphasize the functional relevance of these changes.

## Credit authorship contribution statement

A. Banerjee conducted data analysis and drafted the manuscript, F. Yang contributed to data analysis, J. Dutta provided critical review and feedback, A. Cacciola and M. Hornberger assisted with paper revision and provided technical expertise, M. Saranathan devised the study, supervised data analysis, and contributed to manuscript revisions.

## Supporting information

Supplemental sheet

## Data Availability

All data produced in the present study are available upon reasonable request to the authors

## Acknowledgements

We acknowledge the use of data collected by the Neuroimaging in Frontotemporal Dementia (NIFD/FTLDNI) initiative, supported by the National Institute on Aging (Grant R01 AG032306) and coordinated through the University of California, San Francisco, Memory and Aging Center. FTLDNI data were disseminated by the Laboratory for Neuro Imaging at the University of Southern California. The primary objective of this initiative was to characterize longitudinal clinical and imaging changes in frontotemporal lobar degeneration (FTLD) and evaluate imaging modalities and biomarkers for diagnostic purposes.

This work was additionally supported by the National Institutes of Health - National Institute on Aging (NIH/NIA) Grants R03 AG070750 and R01 AG072669, as well as by the National Institute on Aging Grant R21 AG087392 and the National Institute of Biomedical Imaging and Bioengineering Grant R01 EB032674.

We extend our gratitude to the FTLDNI investigators for their collaborative efforts and commitment to advancing research in neurodegenerative disorders.

## References

1. Bang, J., Spina, S., & Miller, B. L. (2015). Frontotemporal dementia. The Lancet, 386(10004), 1672–1682.

2. Weder, N. D., Aziz, R., Wilkins, K., & Tampi, R. R. (2007). Frontotemporal dementias: a review. Annals of General Psychiatry, 6(1), 1–10.

3. Hornberger, M., Wong, S., Tan, R., Irish, M., Piguet, O., Kril, J., … & Halliday, G. (2012). In vivo and post-mortem memory circuit integrity in frontotemporal dementia and Alzheimer’s disease. Brain, 135(10), 3015–3025.

4. Muñoz-Ruiz, M. Á., Hartikainen, P., Koikkalainen, J., Wolz, R., Julkunen, V., Niskanen, E., … & Soininen, H. (2012). Structural MRI in frontotemporal dementia: Comparisons between hippocampal volumetry, tensor-based morphometry and voxel-based morphometry. PLOS ONE, 7, e52531.

5. Warren, J. D., Rohrer, J. D., & Rossor, M. N. (2013). Frontotemporal dementia. BMJ, 347, f4827.

6. Olney, N. T., Spina, S., & Miller, B. L. (2017). Frontotemporal dementia. Neurologic Clinics, 35(2), 339–374.

7. McKenna, M. C., Lope, J., Bede, P., & Tan, E. L. (2023). Thalamic pathology in frontotemporal dementia: predilection for specific nuclei, phenotype-specific signatures, clinical correlates, and practical relevance. Brain and Behavior, 13(2), e2881.

8. Devenney, E. M., Tse, N. Y., O’Callaghan, C., Kumfor, F., Ahmed, R. M., Caga, J., … & Hodges, J. R. (2024). An attentional and working memory theory of hallucination vulnerability in frontotemporal dementia. Brain Communications, 6(3), fcae123.

9. Sherman, S. M. (2016). Thalamus plays a central role in ongoing cortical functioning. Nature Neuroscience, 19(4), 533–541.

10. Schmahmann, J. D. (2003). Vascular syndromes of the thalamus. Stroke, 34(9), 2264–2278.

11. Segobin, S., Haast, R. A., Kumar, V. J., Lella, A., Alkemade, A., Bach Cuadra, M., … & Hornberger, M. (2024). A roadmap towards standardized neuroimaging approaches for human thalamic nuclei. Nature Reviews Neuroscience, *1*-17.

12. Bocchetta, M., Iglesias, J. E., Neason, M., Cash, D. M., Warren, J. D., & Rohrer, J. D. (2020). Thalamic nuclei in frontotemporal dementia: Mediodorsal nucleus involvement is universal but pulvinar atrophy is unique to C9orf72. Human Brain Mapping, 41(4), 1006–1016.

13. Mohammadi, S., Ghaderi, S., Mohammadi, M., Pashaki, Z. N. A., Khatyal, R., Mohammadian, F., & Mohammadjani, S. (2024). Thalamic alterations in motor neuron diseases: a systematic review of MRI findings. Journal of Integrative Neuroscience, 23(4), 77.

14. Looi, J. C. L., Svensson, L., Lindberg, O., Zandbelt, B. B., Östberg, P., Örndahl, E., & Wahlund, L. O. (2009). Putaminal volume in frontotemporal lobar degeneration and Alzheimer disease: Differential volumes in dementia subtypes and controls. American Journal of Neuroradiology, 30(8), 1552–1560.

15. Looi, J. C., Rajagopalan, P., Walterfang, M., Madsen, S. K., Thompson, P. M., Macfarlane, M. D., … & Velakoulis, D. (2012). Differential putaminal morphology in Huntington’s disease, frontotemporal dementia and Alzheimer’s disease. Australian & New Zealand Journal of Psychiatry, 46(12), 1145–1158.

16. Luo, X., Mao, Q., Shi, J., Wang, X., & Li, C. S. R. (2019). Putamen gray matter volumes in neuropsychiatric and neurodegenerative disorders. World Journal of Psychiatry and Mental Health Research, 3(1).

17. Bocchetta, M., Malpetti, M., Todd, E. G., Rowe, J. B., & Rohrer, J. D. (2021).Looking beneath the surface: The importance of subcortical structures in frontotemporal dementia. Brain Communications, 3(3), fcab158.

18. 18. Möller, C., Dieleman, N., van der Flier, W. M., Versteeg, A., Pijnenburg, Y., Scheltens, P., Barkhof, F., & Vrenken, H. (2024). More atrophy of deep gray matter structures in frontotemporal dementia compared to Alzheimer’s disease. Brain Imaging and Behavior, 18, 66–72.

19. Links, K. A., Chow, T. W., Binns, M., Freedman, M., Stuss, D. T., Scott, C. J. M., Ramirez, J., & Black, S. E. (2018). Apathy is not associated with basal ganglia atrophy in frontotemporal dementia. Journal of Geriatric Psychiatry and Neurology, 23, 34–42.

20. Halabi, C., Halabi, A., Dean, D. L., Wang, P. N., Boxer, A. L., Trojanowski, J. Q., … & Seeley, W. W. (2013). Patterns of striatal degeneration in frontotemporal dementia. Alzheimer Disease & Associated Disorders, 27(1), 74–83.

21. Saranathan, M., Iglehart, C., Monti, M., Tourdias, T., & Rutt, B. (2021). In vivo high-resolution structural MRI-based atlas of human thalamic nuclei. Scientific Data, 8(1), 275.

22. Bernstein, A. S., Rapcsak, S. Z., Hornberger, M., Saranathan, M., & Alzheimer’s Disease Neuroimaging Initiative. (2021). Structural changes in thalamic nuclei across prodromal and clinical Alzheimer’s disease. Journal of Alzheimer’s Disease, 82(1), 361–371.

23. Tourdias, T., Saranathan, M., Levesque, I. R., Su, J., & Rutt, B. K. (2014). Visualization of intra-thalamic nuclei with optimized white-matter-nulled MPRAGE at 7 T. Neuroimage, 84, 534–545.

24. Su, J. H., Thomas, F. T., Kasoff, W. S., Tourdias, T., Choi, E. Y., Rutt, B. K., & Saranathan, M. (2019). Thalamus Optimized Multi Atlas Segmentation (THOMAS): Fast, fully automated segmentation of thalamic nuclei from structural MRI. Neuroimage, 194, 272–282.

25. Vidal, J. P., Danet, L., Péran, P., Pariente, J., Bach Cuadra, M., Zahr, N. M., … & Saranathan, M. (2024). Robust thalamic nuclei segmentation from T1-weighted MRI using polynomial intensity transformation. Brain Structure and Function, 229(5), 1087–1101.

26. Williams, B., Nguyen, D., Vidal, J. P., & Saranathan, M. (2024). Thalamic nuclei segmentation from T1-weighted MRI: Unifying and benchmarking state-of-the-art methods. Imaging Neuroscience, 2, 1–16.

27. Illán-Gala, I., Casaletto, K. B., Borrego-Ecija, S., Arenaza-Urquijo, E. M., Wolf, A., Cobigo, Y., … & Rosen, H. J. (2021). Sex differences in the behavioral variant of frontotemporal dementia: A new window to executive and behavioral reserve. Alzheimer’s & Dementia, 17(8), 1329–1341.

28. Pengo, M., Alberici, A., Libri, I., Benussi, A., Gadola, Y., Ashton, N. J., … & Borroni, B. (2022). Sex influences clinical phenotype in frontotemporal dementia. Neurological Sciences, 43(9), 5281–5287.

29. 29. Overbeek, J. M., Korten, N., Gossink, F., Fieldhouse, J., van de Beek, M., Reus, L., … & Schouws, S. (2020). The value of neuropsychological assessment in the differentiation between behavioral variant frontotemporal dementia and late-onset psychiatric disorders. The Journal of Clinical Psychiatry, 81(1), 12725.

30. Ranasinghe, K. G., Rankin, K. P., Lobach, I. V., Kramer, J. H., Sturm, V. E., Bettcher, B. M., … & Miller, B. L. (2016). Cognition and neuropsychiatry in behavioral variant frontotemporal dementia by disease stage. Neurology, 86(7), 600–610.

31. Bertoux, M., O’Callaghan, C., Flanagan, E., Hodges, J. R., & Hornberger, M. (2015). Fronto-striatal atrophy in behavioral variant frontotemporal dementia and Alzheimer’s disease. Frontiers in Neurology, 6, 147.

32. Macfarlane, M. D., Jakabek, D., Walterfang, M., Vestberg, S., Velakoulis, D., Wilkes, F. A., … & Santillo, A. F. (2015). Striatal atrophy in the behavioural variant of frontotemporal dementia: Correlation with diagnosis, negative symptoms and disease severity. PLOS ONE, 10(6), e0129692.

33. Parnaudeau, S., Bolkan, S. S., & Kellendonk, C. (2018). The mediodorsal thalamus: An essential partner of the prefrontal cortex for cognition. Biological Psychiatry, 83(8), 648–656.

34. Petersen, S. E., Robinson, D. L., & Morris, J. D. (1987). Contributions of the pulvinar to visual spatial attention. Neuropsychologia, 25(1), 97–105.

35. Ogar, J. M., Dronkers, N. F., Brambati, S. M., Miller, B. L., & Gorno-Tempini, M. L. (2007). Progressive nonfluent aphasia and its characteristic motor speech deficits. Alzheimer Disease & Associated Disorders, 21(4), S23–S30.

36. Tan, R. H., Wong, S., Kril, J. J., Piguet, O., Hornberger, M., Hodges, J. R., & Halliday, G. M. (2014). Beyond the temporal pole: Limbic memory circuit in the semantic variant of primary progressive aphasia. Brain, 137(7), 2065–2076.

37. 37. Di Biase, M. A., Tian, Y. E., Bethlehem, R. A., Seidlitz, J., Alexander-Bloch, A. F., Yeo, B. T., & Zalesky, A. (2023). Mapping human brain charts cross-sectionally and longitudinally. Proceedings of the National Academy of Sciences, 120(20), e2216798120.

38. Bethlehem, R. A., Seidlitz, J., White, S. R., Vogel, J. W., Anderson, K. M., Adamson, C., … & Schaare, H. L. (2022). Brain charts for the human lifespan. Nature, 604(7906), 525–533.

39. Benarroch, E. E. (2021). What is the role of the claustrum in cortical function and neurologic disease? Neurology, 96(3), 110–113.

40. Crick, F. C., & Koch, C. (2005). What is the function of the claustrum? Philosophical Transactions of the Royal Society B: Biological Sciences, 360(1458), 1271–1279.

41. Jakabek, D., Power, B. D., Spotorno, N., Macfarlane, M. D., Walterfang, M., Velakoulis, D., … & Santillo, A. F. (2018). Structural and microstructural thalamocortical network disruption in sporadic behavioural variant frontotemporal dementia. NeuroImage: Clinical, 39, 103471.

42. McKenna, M. C., Shing, S. L. H., Murad, A., Lope, J., Hardiman, O., Hutchinson, S., & Bede, P. (2022). Focal thalamus pathology in frontotemporal dementia: Phenotype-associated thalamic profiles. Journal of the Neurological Sciences, 436, 120221.

43. Morel, A., Magnin, M., & Jeanmonod, D. (1997). Multiarchitectonic and stereotactic atlas of the human thalamus. Journal of Comparative Neurology, 387(4), 588–630.

