## Supplemental sheet for "Cross-Sectional and Longitudinal Patterns of Atrophy in Thalamic and Deep Gray Matter Nuclei in Frontotemporal Dementia"

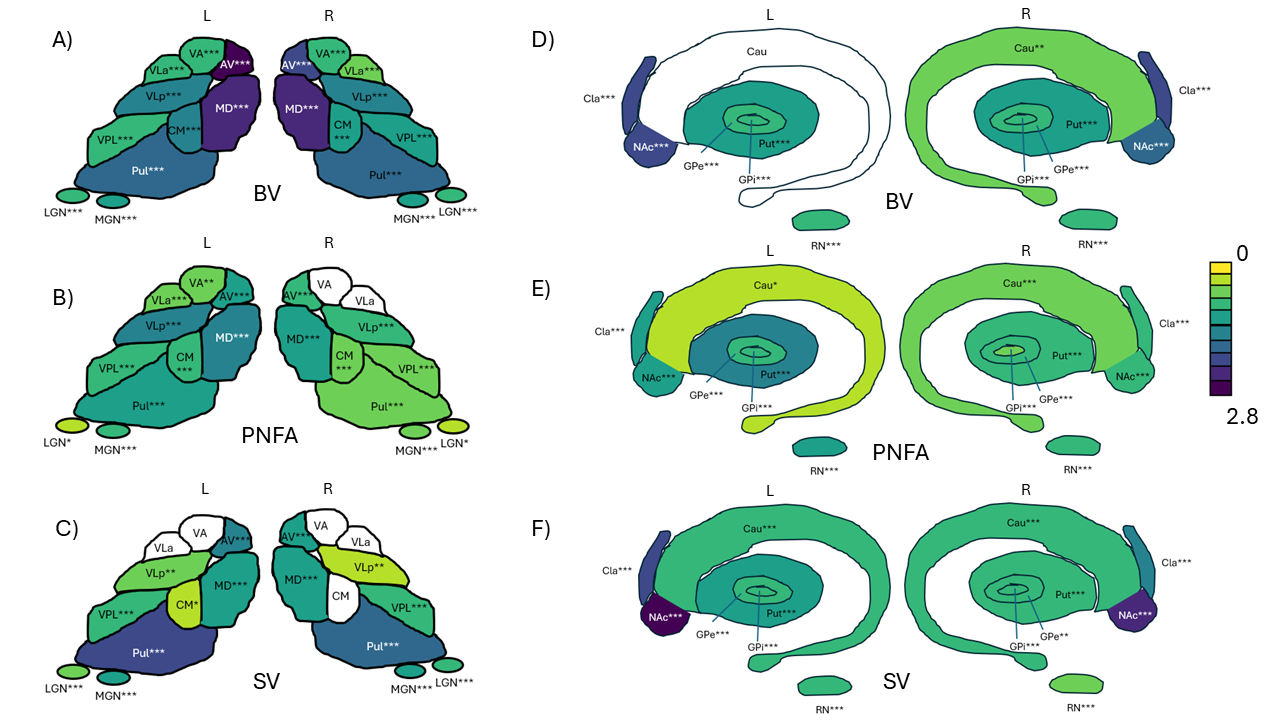

**Supplementary Figure 1. Schematic showing Significant Effect Sizes in Thalamic and other Deep Gray Matter nuclei Regions for Timepoint 2**

Supplementary Figure 1 illustrates significant effect sizes (Cohen’s d) in thalamic and other deep gray matter regions across bvFTD, svFTD, and PNFA. The AV and MD nuclei exhibited the largest effect sizes, highlighting their vulnerability, particularly in bvFTD. Substantial changes were also observed in the pulvinar, putamen, and nucleus accumbens, emphasizing a gradient of atrophy severity, with more pronounced effects in bvFTD and svFTD compared to other regions. These findings underscore the heterogeneity of structural changes across FTD subtypes.

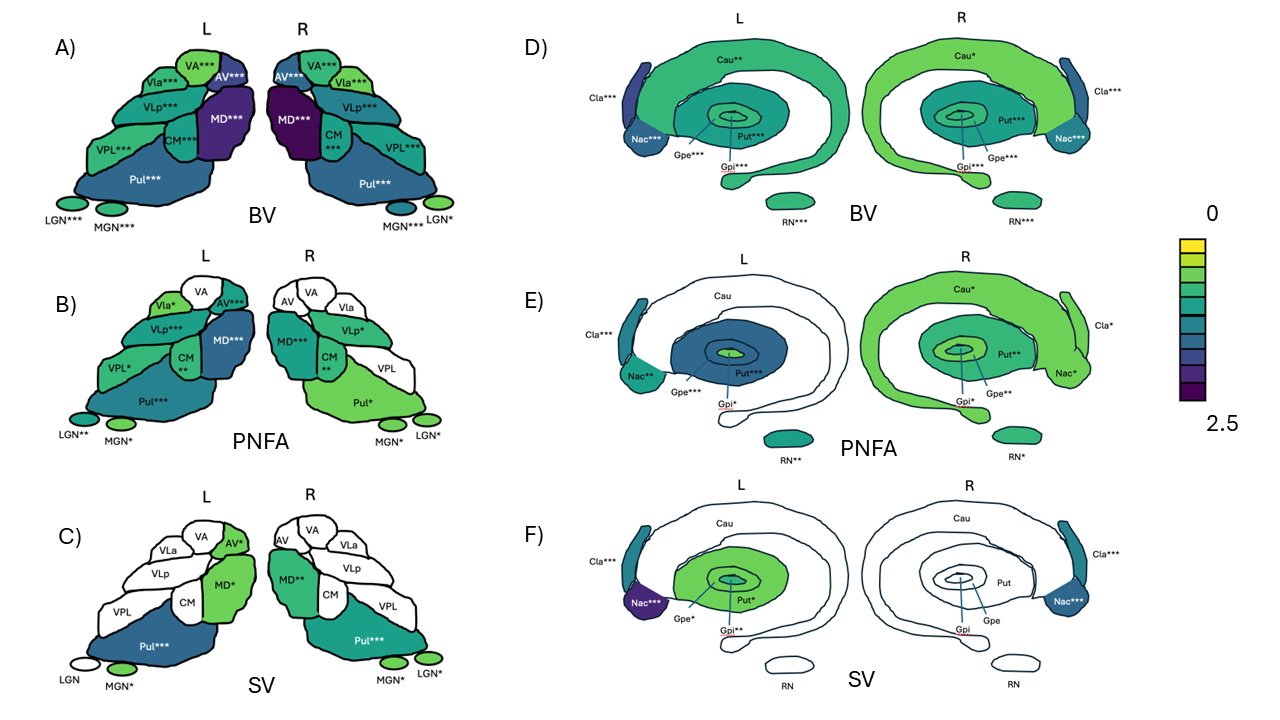

**Supplementary Figure 2. Thalamic and Deep Grey Matter Nuclei Comparisons Between Patient Groups and Controls (Males) in Timepoint 1**

- **A, D:** bvFTD vs. CN, showing significant atrophy in MD, AV, Pul (A) and Cla, NAc, Put (D).
- **B, E:** PNFA vs. CN, highlighting MD, VPL (B) and Cla, NAc (E) alterations.
- **C, F:** svFTD vs. CN, with MD, Pul (C) and Cla, NAc (F) affected.

Color intensity indicates effect size, with significant regions labeled.

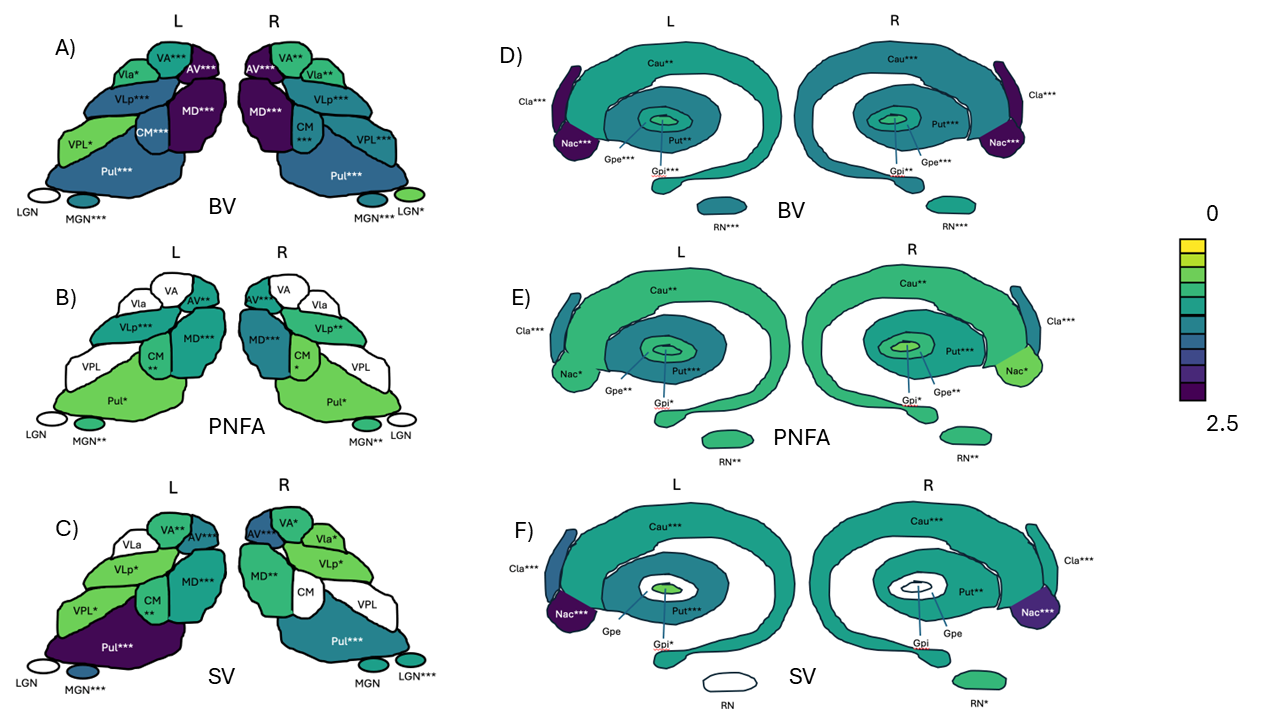

**Supplementary Figure 3. Thalamic and Deep Grey Matter Nuclei Comparisons Between Patient Groups and Controls (Females) in Timepoint 1**

- **A, D:** bvFTD vs. CN, showing significant atrophy in MD, AV, Pul (A) and Cla, NAc, Put (D).
- **B, E:** PNFA vs. CN, highlighting MD, VPL (B) and Cla, NAc (E) alterations.
- **C, F:** svFTD vs. CN, with MD, Pul (C) and Cla, NAc (F) affected.

Color intensity indicates effect size, with significant regions labeled.

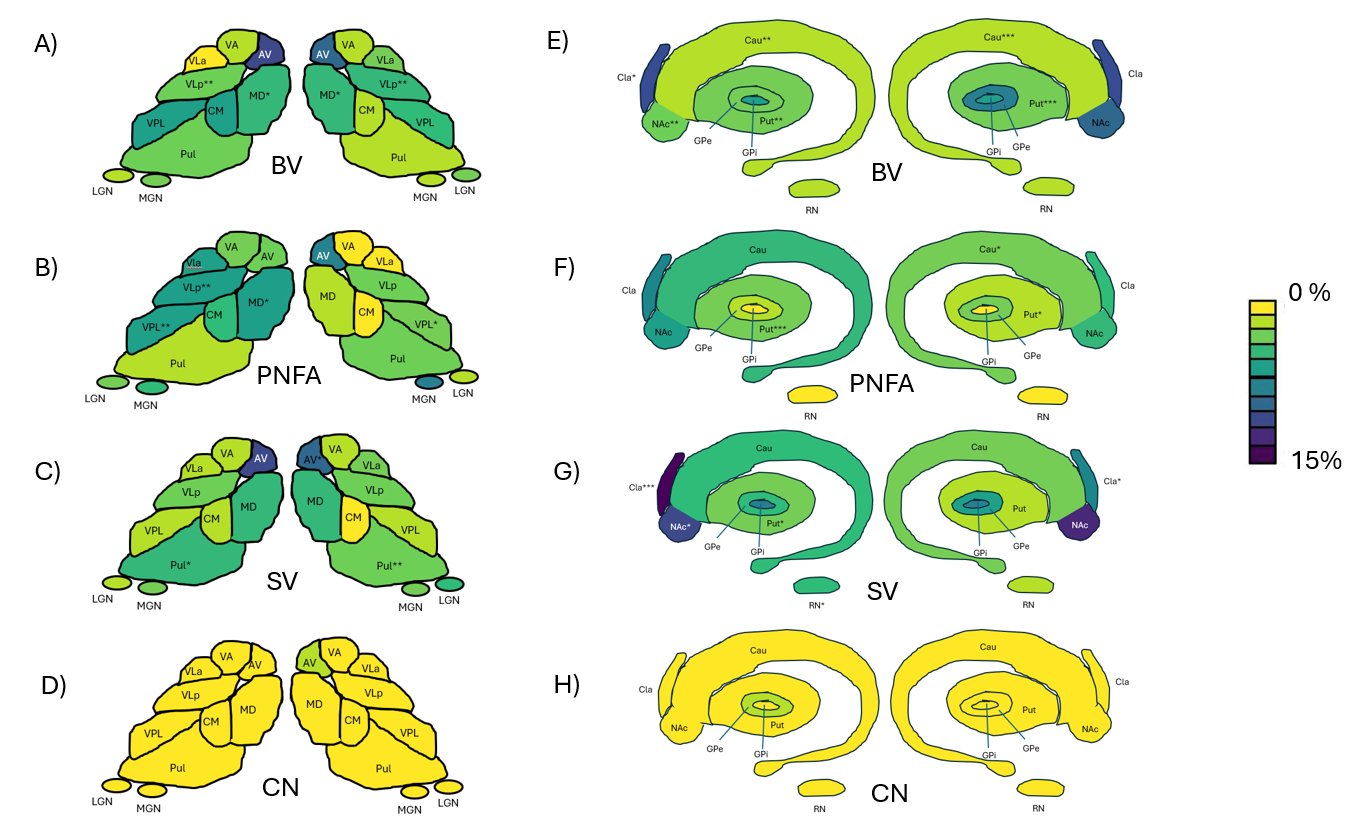

**Supplemental Figure 4: Thalamic and Deep Grey Matter Atrophy in Males Across FTD Subtypes longitudinally**

This figure illustrates the distribution of volume reductions across thalamic and deep grey matter structures in males with bvFTD (A, E), PNFA (B, F), and svFTD (C, G) compared to cognitively normal controls (CN; D, H) from timepoint 1 to timepoint 2.

**Thalamic atrophy (A–D):** MD, VLp, and CM are most affected in bvFTD and PNFA, while AV and Pul show greater reductions in svFTD.

**Other Deep gray matter atrophy (E–H):** bvFTD shows widespread Caudate, Putamen, and Claustrum atrophy, PNFA has localized Putamen loss, and svFTD exhibits pronounced NAc and Claustrum degeneration, indicating subtype-specific vulnerabilities.

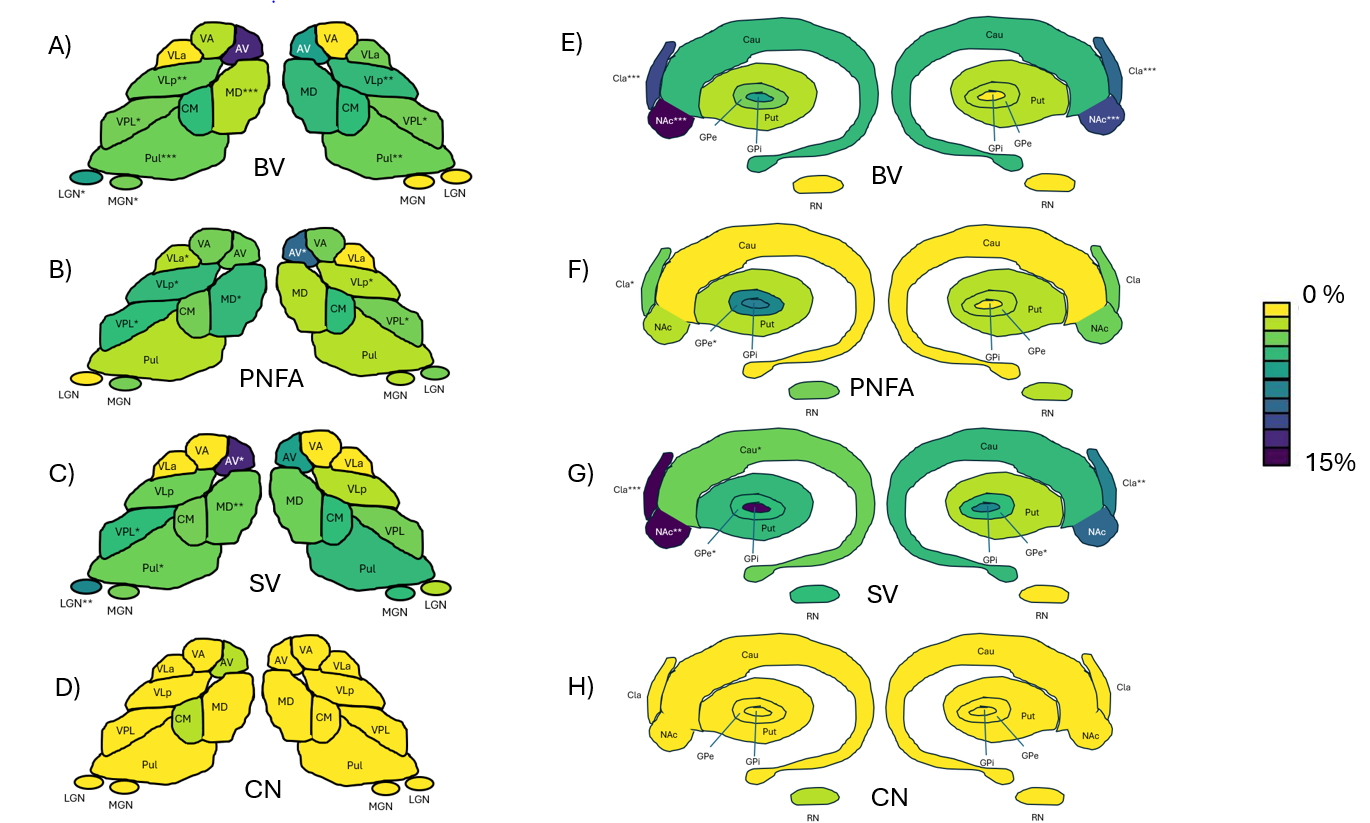

**Supplemental Figure 5: Thalamic and Deep Grey Matter Atrophy in Females Across FTD Subtypes longitudinally**

This figure illustrates volume reductions in thalamic and deep grey matter structures in females with bvFTD (A, E), PNFA (B, F), and svFTD (C, G) compared to cognitively normal controls (CN; D, H) from timepoint 1 to timepoint 2.

**Thalamic atrophy (A–D):** MD, AV, and Pul are most affected, particularly in bvFTD and svFTD. AV shows greater atrophy in svFTD, while MD exhibits widespread involvement across subtypes.

**Other Deep gray matter atrophy (E–H):** NAc and Claustrum show severe atrophy in bvFTD and svFTD, indicating disruptions in motivation and emotional regulation. Unlike males, females exhibit more pronounced NAc degeneration, particularly in svFTD.

| Region | Adjusted mean volumes of CN for TP1 | Adjusted mean volumes of CN for TP2 | Adjusted mean volumes of BV for TP1 | Adjusted mean volumes of BV for TP2 | Adjusted mean volumes of PNFA for TP1 | Adjusted mean volumes of PNFA for TP2 | Adjusted mean volumes of SV for TP1 | Adjusted mean volumes of SV for TP2 |
| --- | --- | --- | --- | --- | --- | --- | --- | --- |
| L WT | 5122.608 | 5040.38 | 4427.891 | 4134.142 | 4607.904 | 4327.766 | 4756.517 | 4485.561 |
| L AV | 98.93 | 93.793 | 57.689 | 41.353 | 79.04 | 70.261 | 79.906 | 63.296 |
| L VA | 263.924 | 259.222 | 244.413 | 232.486 | 251.743 | 240.09 | 256.023 | 252.229 |
| L VLa | 79.889 | 78.996 | 73.187 | 69.973 | 75.76 | 71.081 | 79.928 | 77.705 |
| L VLp | 785.208 | 775.504 | 689.658 | 640.483 | 692.74 | 646.526 | 770.525 | 723.813 |
| L VPL | 295.618 | 291.745 | 275.084 | 257.165 | 276.129 | 256.215 | 282.526 | 259.768 |
| L Pul | 1197.412 | 1168.494 | 1014.874 | 936.454 | 1084.57 | 1013.63 | 989.676 | 903.958 |
| L LGN | 97.658 | 95.621 | 87.296 | 79.029 | 88.857 | 87.955 | 91.495 | 81.111 |
| L MGN | 60.522 | 59.619 | 53.114 | 50.104 | 55.567 | 52.452 | 53.978 | 50.17 |
| L CM | 98.422 | 94.35 | 81.525 | 73.969 | 85.769 | 80.026 | 93.854 | 87.655 |
| L MD | 600.993 | 593.727 | 455.371 | 403.613 | 511.25 | 450.665 | 547.811 | 487.678 |
| L Nac | 612.004 | 595.59 | 432.98 | 392.119 | 529.709 | 479.195 | 392.532 | 310.283 |
| L Cau | 3342.223 | 3308.271 | 2985.259 | 3153.203 | 3084.055 | 3073.108 | 3042.781 | 2903.368 |
| L Cla | 848.237 | 830.563 | 569.235 | 496.762 | 673.911 | 597.478 | 643.188 | 500.967 |
| L GPe | 353.226 | 345.159 | 296.628 | 271.258 | 287.335 | 277.779 | 320.423 | 279.502 |
| L GPi | 154.611 | 150.23 | 117.903 | 99.089 | 124.801 | 108.778 | 122.111 | 103.357 |
| L Put | 4695.596 | 4652.954 | 4183.704 | 3995.877 | 4065.407 | 3912.914 | 4258.872 | 4010.467 |
| L RN | 195.424 | 190.59 | 167.605 | 159.022 | 169.623 | 164.627 | 183.779 | 161.679 |
| R WT | 5171.101 | 5085.643 | 4470.352 | 4224.762 | 4855.656 | 4647.16 | 4880.3 | 4628.473 |
| R AV | 100.278 | 94.898 | 61.346 | 48.92 | 84.749 | 74.578 | 84.494 | 66.634 |
| R VA | 276.817 | 273.678 | 256.578 | 245.71 | 272.088 | 264.971 | 270.701 | 265.268 |
| R VLa | 82.047 | 81.173 | 74.614 | 72.456 | 78.113 | 77.486 | 80.869 | 79.081 |
| R VLp | 814.175 | 804.44 | 705.673 | 661.641 | 746.553 | 724.524 | 789.777 | 755.251 |
| R VPL | 283.38 | 279.307 | 255.992 | 238.887 | 270.376 | 254.263 | 275.176 | 253.509 |
| R Pul | 1195.366 | 1166.626 | 1004.671 | 948.284 | 1122.254 | 1077.058 | 1060.533 | 960.033 |
| R LGN | 96.596 | 95.802 | 87.578 | 80.908 | 93.013 | 87.543 | 85.085 | 77.051 |
| R MGN | 62.417 | 61.084 | 53.217 | 51.324 | 57.516 | 55.623 | 58.293 | 52.218 |
| R CM | 95.961 | 92.668 | 79.815 | 73.544 | 85.328 | 80.84 | 91.998 | 87.285 |
| R MD | 614.991 | 600.123 | 450.825 | 422.347 | 537.993 | 501.011 | 550.187 | 510.357 |
| R Nac | 558.926 | 546.109 | 396.632 | 365.192 | 501.752 | 454.068 | 391.982 | 314.934 |
| R Cau | 3445.87 | 3418.433 | 3117.927 | 3147.925 | 3135.936 | 3098.8 | 3167.816 | 3067.009 |
| R Cla | 855.439 | 827.373 | 590.528 | 499.324 | 732.06 | 664.158 | 678.192 | 569.129 |
| R GPe | 332.505 | 326.15 | 282.169 | 260.568 | 290.507 | 267.08 | 312.429 | 270.73 |
| R GPi | 152.583 | 151.246 | 113.793 | 103.455 | 122.956 | 117.062 | 136.456 | 107.632 |
| R Put | 4653.633 | 4609.854 | 4150.195 | 3969.083 | 4241.314 | 4144.508 | 4367.073 | 4135.981 |
| R RN | 190.186 | 189.816 | 165.248 | 157.532 | 165.294 | 159.298 | 172.186 | 164.324 |

**Supplementary Table 1. Adjusted Mean Volumes (in mm³) of Thalamic Nuclei and other Deep Grey Matter Nuclei Across Groups at Two Time Points**

**Description**: Supplementary Table 1 presents the adjusted mean values for different thalamic nuclei and related brain regions across four groups: CN, bvFTD, PNFA, svFTD at two distinct time points (TP1 and TP2). The data includes values for the left (L) and right (R) hemispheres for each region. Adjusted means were computed using statistical models that account for confounding variables such as age, sex, and ICV. This comparison highlights temporal dynamics and group-specific changes in structural volumes, providing insights into disease-specific progression patterns in neurodegenerative disorders.

| Regions | Cohen’s D for bvFTD vs CN at TP2 | Percent reduction of adjusted mean in bvFTD vs CN at TP2 | Cohen’s D for PNFA vs CN at TP2 | Percent reduction of adjusted mean in PNFA vs CN at TP2 | Cohen’s D for svFTD vs CN at TP2 | Percent reduction of adjusted mean in svFTD vs CN at TP2 |
| --- | --- | --- | --- | --- | --- | --- |
| L WT | 2.137*** | 17.98 | 1.68*** | 14.138 | 1.308*** | 11.007 |
| L AV | 2.583*** | 55.91 | 1.159*** | 25.089 | 1.502*** | 32.515 |
| L VA | 0.934*** | 10.314 | 0.668** | 7.38 | 0.244 | 2.697 |
| L VLa | 0.935*** | 11.422 | 0.82*** | 10.019 | 0.134 | 1.634 |
| L VLp | 1.566*** | 17.411 | 1.496*** | 16.632 | 0.6** | 6.666 |
| L VPL | 0.999*** | 11.853 | 1.027*** | 12.178 | 0.924*** | 10.96 |
| L Pul | 1.744*** | 19.858 | 1.164*** | 13.253 | 1.988*** | 22.639 |
| L LGN | 0.94*** | 17.352 | 0.435* | 8.018 | 0.822*** | 15.175 |
| L MGN | 1.391*** | 15.959 | 1.048*** | 12.021 | 1.382*** | 15.848 |
| L CM | 1.437*** | 21.602 | 1.01*** | 15.181 | 0.472* | 7.096 |
| L MD | 2.381*** | 32.02 | 1.792*** | 24.096 | 1.328*** | 17.862 |
| L Nac | 1.974*** | 34.163 | 1.129*** | 19.543 | 2.769*** | 47.903 |
| L Cau | 0.367 | 4.687 | 0.557* | 7.108 | 0.959*** | 12.239 |
| L Cla | 2.181*** | 40.19 | 1.523*** | 28.064 | 2.154*** | 39.683 |
| L GPe | 1.086*** | 21.411 | 0.99*** | 19.522 | 0.965*** | 19.022 |
| L GPi | 1.105*** | 34.042 | 0.895*** | 27.593 | 1.013*** | 31.201 |
| L Put | 1.282*** | 14.122 | 1.444*** | 15.905 | 1.254*** | 13.808 |
| L RN | 1.015*** | 16.563 | 0.834*** | 13.622 | 0.929*** | 15.169 |
| R WT | 2.188*** | 16.928 | 1.114*** | 8.622 | 1.162*** | 8.989 |
| R AV | 2.1*** | 48.451 | 0.928*** | 21.413 | 1.291*** | 29.783 |
| R VA | 0.95*** | 10.219 | 0.296 | 3.182 | 0.286 | 3.073 |
| R VLa | 0.827*** | 10.739 | 0.35 | 4.542 | 0.198 | 2.577 |
| R VLp | 1.6*** | 17.751 | 0.895*** | 9.934 | 0.551** | 6.115 |
| R VPL | 1.347*** | 14.472 | 0.834*** | 8.966 | 0.86*** | 9.236 |
| R Pul | 1.81*** | 18.716 | 0.742*** | 7.677 | 1.712*** | 17.709 |
| R LGN | 0.857*** | 15.547 | 0.475* | 8.62 | 1.078*** | 19.572 |
| R MGN | 1.338*** | 15.979 | 0.749*** | 8.94 | 1.215*** | 14.514 |
| R CM | 1.291*** | 20.638 | 0.799*** | 12.764 | 0.363 | 5.809 |
| R MD | 2.459*** | 29.623 | 1.371*** | 16.515 | 1.242*** | 14.958 |
| R Nac | 1.754*** | 33.128 | 0.892*** | 16.854 | 2.241*** | 42.331 |
| R Cau | 0.646** | 7.913 | 0.764*** | 9.35 | 0.84*** | 10.28 |
| R Cla | 2.049*** | 39.649 | 1.019*** | 19.727 | 1.613*** | 31.213 |
| R GPe | 1.046*** | 20.108 | 0.942*** | 18.111 | 0.884*** | 16.992 |
| R GPi | 0.962*** | 31.598 | 0.688*** | 22.602 | 0.878*** | 28.837 |
| R Put | 1.281*** | 13.9 | 0.931*** | 10.095 | 0.948*** | 10.28 |
| R RN | 1.014*** | 17.008 | 0.959*** | 16.078 | 0.801*** | 13.43 |

**Supplementary Table 2. Effect Sizes (Cohen’s D) and Percentage Reduction in Mean Volume for bvFTD, PNFA, and svFTD Compared to Controls (CN) at Timepoint 2 (TP2).**

**Description:** Supplementary Table 2 presents the effect sizes (Cohen’s D) and percentage reductions in mean volumes for bvFTD, PNFA, and svFTD compared to the control group (CN) at timepoint 2 (TP2). Cohen’s D values indicate the magnitude of atrophy in deep gray matter nuclei across FTD subtypes, with higher values reflecting greater group differences. Statistically significant results are denoted by asterisks (*p < 0.05, **p < 0.01, ***p < 0.001).

| Neuropsych Test | Region | Correlation (r) and Significance | FTD Subtype |
| --- | --- | --- | --- |
| BNTCORR | Left AV | 0.257* | bvFTD |
| BNTCORR | Left VLp | 0.279* | bvFTD |
| BNTCORR | Left Mediodorsal | 0.283* | bvFTD |
| BNTCORR | Left Accumbens | 0.339** | bvFTD |
| PPVTVRB | Left Mediodorsal | 0.362* | bvFTD |
| PPVTVRB | Left Claustrum | 0.361* | bvFTD |
| PPVTVRB | Right Mediodorsal | 0.315* | bvFTD |
| PPVTVRB | Right Red Nucleus | 0.304* | bvFTD |
| PPVTANI | Left Claustrum | 0.393** | bvFTD |
| PPVTANI | Right Mediodorsal | 0.324* | bvFTD |
| DIGITBW | Left AV | 0.353** | bvFTD |
| DIGITBW | Left VLp | 0.399** | bvFTD |
| DIGITBW | Left Centromedian | 0.263* | bvFTD |
| DIGITBW | Left Mediodorsal | 0.308* | bvFTD |
| DIGITBW | Left Putamen | 0.279* | bvFTD |
| DIGITBW | Left Red Nucleus | 0.365** | bvFTD |
| DIGITBW | Right VLp | 0.406*** | bvFTD |
| DIGITBW | Right Centromedian | 0.333** | bvFTD |
| DIGITBW | Right Mediodorsal | 0.263* | bvFTD |
| DIGITBW | Right GPe | 0.356** | bvFTD |
| DIGITBW | Right Putamen | 0.266* | bvFTD |
| BNTCORR | Left Pulvinar | 0.378* | PNFA |
| BNTCORR | Left Centromedian | 0.369* | PNFA |
| BNTCORR | Right Pulvinar | 0.39* | PNFA |
| PPVTVRB | Left VLp | 0.436* | PNFA |
| PPVTVRB | Left Pulvinar | 0.455* | PNFA |
| PPVTVRB | Left Mediodorsal | 0.598*** | PNFA |
| PPVTVRB | Left Claustrum | 0.504** | PNFA |
| PPVTVRB | Left Putamen | 0.386* | PNFA |
| PPVTVRB | Right AV | 0.384* | PNFA |
| PPVTVRB | Right Accumbens | 0.482** | PNFA |
| PPVTVRB | Right Claustrum | 0.397* | PNFA |
| PPVTDES | Left Mediodorsal | 0.472** | PNFA |
| PPVTDES | Left Claustrum | 0.403* | PNFA |
| PPVTDES | Left Putamen | 0.369* | PNFA |
| PPVTDES | Right Claustrum | 0.503** | PNFA |
| PPVTANI | Left Pulvinar | 0.374* | PNFA |
| PPVTANI | Left Mediodorsal | 0.513** | PNFA |
| PPVTANI | Left Claustrum | 0.467** | PNFA |
| PPVTANI | Left Putamen | 0.388* | PNFA |
| PPVTANI | Right Mediodorsal | 0.401* | PNFA |
| PPVTANI | Right Accumbens | 0.406* | PNFA |
| PPVTANI | Right Claustrum | 0.508** | PNFA |
| PPVTINA | Left VLp | 0.398* | PNFA |
| PPVTINA | Left Mediodorsal | 0.602*** | PNFA |
| PPVTINA | Left Claustrum | 0.436* | PNFA |
| PPVTINA | Right Mediodorsal | 0.422* | PNFA |
| PPVTINA | Right Claustrum | 0.387* | PNFA |
| DIGITFW | Left Pulvinar | 0.398* | PNFA |
| DIGITFW | Left Centromedian | 0.364* | PNFA |
| DIGITFW | Right Putamen | 0.349* | PNFA |
| DIGITBW | Left Pulvinar | 0.354* | PNFA |
| DIGITBW | Left Centromedian | 0.374* | PNFA |
| DIGITBW | Left Gpi | 0.378* | PNFA |
| MTCORR | Left AV | 0.355* | PNFA |
| MTCORR | Left VLp | 0.381* | PNFA |
| MTCORR | Left Pulvinar | 0.359* | PNFA |
| MTCORR | Left Mediodorsal | 0.38* | PNFA |
| MTCORR | Left Putamen | 0.364* | PNFA |
| MTCORR | Right VLp | 0.416* | PNFA |
| MTCORR | Right Accumbens | 0.379* | PNFA |

**Supplementary Table 3. Pearson Correlations Between Neuropsychological Tests and Brain Regions Across FTD Subtypes**

**Description**: Supplementary Table 3 presents Pearson correlation coefficients between neuropsychological test scores and structural changes in thalamic and subcortical regions across bvFTD and PNFA subtypes. Significant correlations were identified in bvFTD, including BNTCORR with the left accumbens and mediodorsal nucleus, and DIGITBW with the right VLp and centromedian nuclei. In PNFA, strong correlations were observed for PPVTVRB with the left mediodorsal nucleus and left claustrum, and PPVTANI with the left pulvinar and right accumbens. The highest correlation was noted for PPVTINA with the left mediodorsal nucleus in PNFA. These findings underscore the importance of the mediodorsal nucleus, pulvinar, and accumbens in neuropsychological performance. No significant correlations were observed in svFTD, highlighting a lack of structural-functional associations for this subtype within the tested regions.
